# High-level clarithromycin resistance: a metabolic vulnerability exploited by bismuth in *Helicobacter pylori*

**DOI:** 10.64898/2026.04.29.26351907

**Authors:** Chunmeng He, Ying Huang

**Affiliations:** Department of Gastroenterology, Children’s Hospital of Fudan University, National Children’s Medical Center, Shanghai, China

**Keywords:** *Helicobacter pylori*, Antibiotic Resistance, Clarithromycin Resistance, Bismuth-based Therapy, Iron Metabolism

## Abstract

**Background & Aims:** Clarithromycin (CLA) resistance severely compromises the efficacy of triple therapy (TT) against *Helicobacter pylori* (*H. pylori*). Bismuth-based regimens exhibit greater efficacy against CLA-resistant *H. pylori* than against strains resistant to other antibiotics, suggesting a resistance-specific vulnerability rather than broad antimicrobial activity. The mechanistic basis for this selectivity, however, remains unknown. We hypothesized that high-level CLA resistance confers a metabolically targetable vulnerability that can be exploited by bismuth, and that a quantitative MIC of CLA threshold could identify this responsive subset.

**Methods:** We conducted a real-world retrospective analysis of 4,610 pediatric patients with *H. pylori* infection treated between 2019 and 2024, among whom 1,844 (40%) had complete follow-up data for eradication assessment. In parallel, we prospectively enrolled 51 patients with culture-positive isolates—the largest liquid checkerboard panel reported to date—to evaluate bismuth–CLA interactions and track treatment outcomes. Mechanistic validation included transcriptomic profiling and functional assays of iron and ATP metabolism, with iron chelation and supplementation experiments.

**Results:** In the retrospective real-world cohort (n = 4,610; 1,844 with follow-up), bismuth quadruple therapy (BQT) achieved superior eradication specifically in CLA-resistant infections (93.1% vs 68.8% with TT; *p* = 0.017). *In vitro*, bismuth–CLA synergy was exclusive to resistant strains and intensified with increasing MIC of CLA. Mechanistically, bismuth triggered coordinated depletion of intracellular iron and ATP—a phenotype mimicked by iron chelation and reversed by iron supplementation. A baseline MIC of CLA ≥16 μg/mL robustly predicted this synergy (AUC = 0.991) and was prospectively validated in an independent patient subset: bismuth cured 96% of high-level resistant patients (MIC ≥ 16 μg/mL) versus 0% with triple therapy (*p* < 0.001).

**Conclusion:** High-level CLA resistance defines an iron-dependent metabolic vulnerability in *H. pylori* that is selectively targeted by bismuth. The MIC threshold of ≥ 16 μg/mL provides the first clinically actionable biomarker for resistance-guided therapy, transforming a marker of treatment failure into a positive predictor of bismuth response. These findings establish the mechanistic and clinical foundation for MIC-stratified eradication strategies and inform future randomized trials aimed at precision management of antibiotic-resistant *H. pylori* infection.

**Graphical abstract:** *Left:* High-level clarithromycin (CLA) resistance defines a distinct physiological phenotype in *Helicobacter pylori*, in which an elevated MIC of CLA (≥ 16 µg/mL) predicts poor eradication with triple therapy (TT) but favorable response to bismuth-containing quadruple therapy (BQT).

*Middle:* Mechanistically, CLA resistance is associated with upregulation of the ferric uptake regulator Fur, leading to reprogrammed iron homeostasis and an increased metabolic burden. Colloidal bismuth subcitrate (CBS) disrupts Fur-dependent iron regulation, exacerbates iron-restricted metabolic stress, and compromises cellular integrity, thereby selectively sensitizing CLA-resistant bacteria to antibiotic killing.

*Right:* Translational implication of reframing antibiotic resistance as a therapeutic vulnerability—bismuth-based regimens function as a “key” that unlocks resistance-associated metabolic liabilities, delays resistance evolution, and improves treatment outcomes. 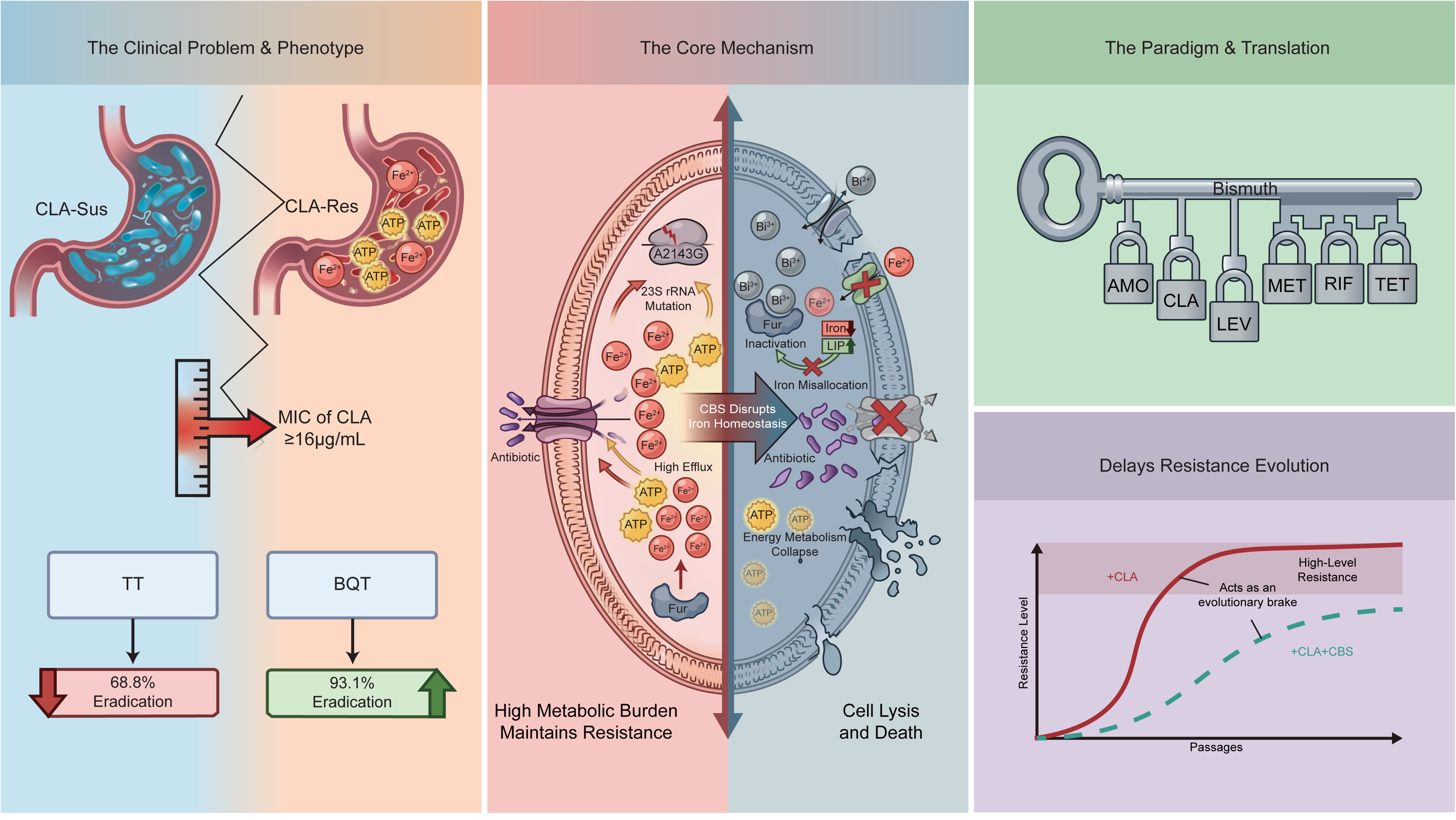

## Introduction

The escalating global prevalence of clarithromycin (CLA) resistance has critically undermined the efficacy of standard triple therapy (TT) for *Helicobacter pylori* (*H. pylori*) eradication, positioning it as a primary driver of treatment failure and a persistent risk factor for gastric cancer.^1–3^ In response, international consensus guidelines now endorse bismuth-containing quadruple therapy (BQT) as a preferred first-line or rescue option, even in regions with high CLA resistance.^4–10^ This recommendation is robustly supported by meta-analytic data. For instance, a pooled analysis of patients with CLA-resistant strains demonstrated a dramatic therapeutic advantage for bismuth-containing regimens over non-bismuth therapies (pooled eradication rate: 86.7% vs 33.3%; OR = 10.64, 95% CI: 2.96 - 39.53).^11^ Despite this compelling clinical evidence, clinicians lack quantitative, MIC-based criteria to prospectively identify which patients with CLA-resistant infections are most likely to derive such profound benefit from bismuth, limiting the precision of treatment selection in routine practice. Bismuth consistently eradicates CLA-resistant *H. pylori* in clinical practice, yet whether this efficacy stems from targeting a vulnerability intrinsic to the resistant state itself has not been explored.^11–15^ Bismuth salts possess broad antimicrobial activity, yet their selective and durable efficacy against CLA-resistant *H. pylori* suggests mechanisms beyond mere nonspecific bactericidal effects.^16–22^ At the molecular level, bismuth is known to interact with the ferric uptake regulator (Fur), a master transcriptional regulator of bacterial iron homeostasis and oxidative stress responses.^23^ Concurrently, high-level CLA resistance in *H. pylori*, often mediated by 23S rRNA point mutations, is associated with broader physiological adaptations, including altered membrane permeability, efflux pump activity, and metabolic stress.^15, 24^ Whether these resistance-associated adaptations create a distinct physiological state—a specific vulnerability—that modulates susceptibility to bismuth remains largely unexplored.

This gap raises a clinically testable and biologically significant question: Does high-level CLA resistance in *H. pylori* represent not merely a marker of antibiotic failure, but a distinct biological state that can be therapeutically exploited with bismuth? Emerging evidence indicates that antibiotic resistance frequently imposes fitness costs and metabolic reprogramming, which can alter bacterial susceptibility to non-antibiotic agents, including metal-based compounds.^25^ Given the established link between bismuth and Fur-mediated metal regulation, perturbations in iron homeostasis could plausibly provide a mechanistic context linking the physiology of CLA resistance to selective bismuth responsiveness.^26, 27^ Recent advances in the field of collateral sensitivity have established that antibiotic resistance can create exploitable metabolic vulnerabilities.^25^ Here, we demonstrate that high-level CLA resistance in *H. pylori* defines such a vulnerability—iron dyshomeostasis—that is selectively targeted by bismuth, providing the first clinical validation of this concept in a major gastrointestinal pathogen. The dramatic clinical differential observed in resistant populations^11^ suggests that the magnitude of this effect may be quantitatively linked to the level of resistance itself.

To address this hypothesis, we adopted an integrated clinical–experimental approach, combining a retrospective cohort analysis, a prospective validation study, and mechanistic investigations in clinical isolates. Our goal was to determine whether quantitative MIC values can identify a metabolically vulnerable, bismuth-responsive bacterial subset—providing a mechanistic and translational basis for MIC-guided precision therapy.

## Methods

### 1. Study Design and Overview

This study employed an integrative clinical–experimental design to determine whether quantitative levels of CLA resistance identify *H. pylori* infections that preferentially benefit from BQT. The work comprised two distinct components: (1) a retrospective, real-world cohort analysis of eradication outcomes using historical patient data (2019–2024), and (2) a prospective observational cohort study (May–August 2024) in which *H. pylori* isolates were collected during routine care and subjected to phenotypic, molecular, and functional analyses. All treatment decisions were made independently by attending physicians according to clinical guidelines; no interventional allocation was performed by the study protocol.

### 2. Clinical Cohorts and Data Collection

#### 2.1. Retrospective cohort

We conducted a single-center study at Children’s Hospital of Fudan University including patients aged 6–18 years with confirmed *H. pylori* infection and no prior eradication therapy. Demographic data, eradication regimens, treatment outcomes, and antimicrobial susceptibility results were extracted from electronic medical records. Eradication success was defined as a negative test ≥ 4 weeks after therapy completion.

#### 2.2. Prospective validation cohort

Between May and August 2024, patients undergoing clinically indicated endoscopy with culture-positive *H. pylori* were enrolled for MIC determination and treatment outcome follow-up.

### 3. Antimicrobial Susceptibility and Synergy Testing

MICs for CLA and other antibiotics were determined by agar dilution. CLA resistance was defined per EUCAST criteria (MIC > 0.25 μg/mL). Checkerboard assays were performed to assess interactions between colloidal bismuth subcitrate (CBS) and antibiotics; synergy was defined as fractional inhibitory concentration index (FICI) ≤ 0.5.

### 4. Mechanistic Studies

In selected clinical isolates, we performed: (1) 23S rRNA sequencing for resistance-associated mutations; (2) transcriptomic profiling (RNA-seq) to identify CBS-affected pathways; (3) quantification of intracellular iron (total and labile pools) and ATP levels; (4) real-time ethidium bromide (EtBr) efflux assays to assess efflux pump activity; and (5) serial passage experiments to evaluate CBS effects on resistance emergence. Iron availability was experimentally manipulated using deferoxamine (DFOM) or ferric chloride (FeCl_3_) supplementation.

### 5. Statistical Analysis

Analyses were performed using SPSS 20.0, GraphPad Prism 9.0, and R 4.5.2. Continuous variables were compared using Student’s t-test or one-way ANOVA; categorical variables using χ² or Fisher’s exact tests. Receiver operating characteristic (ROC) analysis identified MIC thresholds predictive of CBS–CLA synergy. All tests were two-tailed, with *p* < 0.05 considered statistically significant.

### 6. Supplementary Methods

Detailed experimental procedures and data are available in the Supplementary Methods.

## Results

### 1. Real-world evidence reveals a selective eradication advantage of bismuth-containing quadruple therapy in CLA-resistant *H. pylori*

Using real-world clinical data, we performed a retrospective analysis of 4,610 patients with *H. pylori* infection treated between 2019 and 2024, of whom 1,844 (40.0%) completed follow-up. BQT achieved the highest eradication rate (87.1%, 867/995; 95% CI 84.9% – 89.2%), significantly exceeding those of concomitant therapy (CT) (81.8%, 36/44; 95% CI 67.3% – 91.8%) and TT (72.9%, 587/805; 95% CI 69.7% – 76.0%; χ^2^ = 58.0, *p* < 0.0001; Table S2).

This efficacy advantage occurred against a background of high antimicrobial resistance in the study population. Among 743 cultured isolates, 50.1% (372/743) were resistant to at least one antibiotic. Among the tested antibiotics, resistance was most prevalent to metronidazole (MET, 89.0%), followed by CLA (41.7%), whereas resistance to amoxicillin (AMO) and levofloxacin (LEV) remained uncommon (4.3% and 1.1%, respectively; Table S1).

Among 1,844 completed follow-up patients, *H. pylori* was successfully cultured from 313 patients (29.2% of those with follow-up), and antibiotic susceptibility testing was performed for 176 isolates (Figure S1). Culture was not performed in all patients due to selective clinical sampling, as detailed in the Methods section. Subgroup analyses showed that eradication rates did not differ significantly among the three treatment regimens in resistance-stratified comparisons, likely reflecting the very limited number of patients receiving CT (Table S3). However, in pairwise analysis, BQT achieved a significantly higher eradication rate than TT in CLA-resistant infections (93.1% [27/29] vs 68.8% [22/32]; χ^2^ = 5.71, *p* = 0.017; Figure 1a). In contrast, no significant difference was observed between these two regimens in MET-resistant infections (80.0% vs 73.8%, *p* = 0.367).

**Figure 1.**
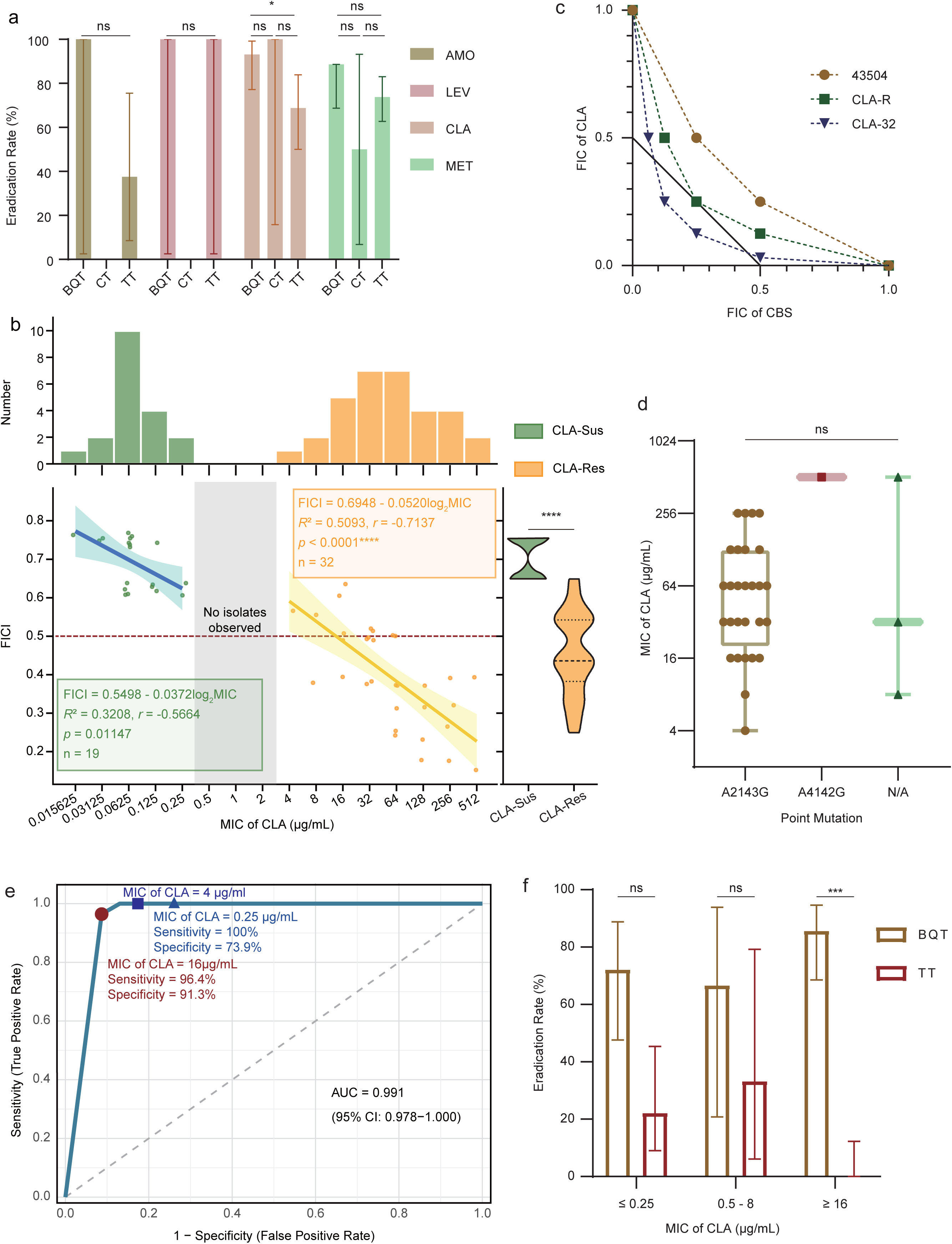
Clarithromycin (CLA) resistance is associated with enhanced responsiveness to colloidal bismuth subcitrate (CBS). **(a)** Eradication rates of different therapeutic regimens containing amoxicillin (AMO), levofloxacin (LEV), CLA, or metronidazole (MET) under bismuth-containing quadruple therapy (BQT), concomitant therapy (CT), or triple therapy (TT). Data are presented as mean ± SD. Statistical significance was assessed using Student’s t-test. ns, not significant; \**p* < 0.05. **(b)** Distribution of baseline CLA minimum inhibitory concentrations (MICs) among CLA-susceptible (CLA-Sus) and CLA-resistant (CLA-Res) *H. pylori* clinical isolates. Upper panels show the bimodal MIC distribution, segregating isolates into clearly susceptible and resistant populations with no intermediate phenotypes. Lower panels depict the relationship between baseline MIC of CLA and the fractional inhibitory concentration index (FICI) of the CBS–CLA combination. Each dot represents an individual isolate. Dashed red lines indicate the threshold for synergy (FICI ≤ 0.5). Solid lines represent linear regression fits with shaded areas indicating 95% confidence intervals. Regression analyses were performed separately for CLA-susceptible and CLA-resistant groups. **(c)** Isobologram analysis of the interaction between CLA and CBS based on checkerboard microdilution assays. Fractional inhibitory concentrations (FICs) of CLA are plotted against those of CBS, with each point representing an individual strain. The dashed diagonal line represents the line of additivity (FIC of CLA + FIC of CBS = 1). Data points falling below the line indicate synergistic interactions. **(d)** Distribution of MICs of CLA stratified by 23S rRNA mutation status (A2143G, A2142G, or no mutation detected). Data are shown as median with interquartile range. **(e)** Receiver operating characteristic (ROC) curve assessing the performance of MIC of CLA in predicting CBS–CLA synergy. The area under the curve (AUC) with the 95% confidence interval is shown. Sensitivity and specificity correspond to the optimal MIC cut-off (MIC of CLA ≥ 16 µg/mL) determined by Youden’s index. For reference, the operational resistance threshold used in this study (MIC of CLA = 4 µg/mL), as well as the clinical resistance breakpoints defined by EUCAST (MIC of CLA > 0.25 µg/mL), are indicated on the ROC curve. **(f)** Eradication rates of BQT versus TT stratified by baseline MIC of CLA. Data are from the prospective validation cohort of 49 paediatric patients (MIC of CLA ≤ 0.25 μg/mL: n = 13; MIC of CLA 0.5–8 μg/mL: n = 11; MIC of CLA ≥ 16 μg/mL: n = 25). Bars represent mean ± 95% confidence interval. \*\*\**p* < 0.001 by Fisher’s exact test (BQT vs TT in MIC of CLA ≥ 16 group); ns, not significant. The MIC of CLA threshold of ≥ 16 μg/mL was pre-specified based on mechanistic synergy data and validated as predictive of therapeutic response.

### 2. Bismuth–CLA synergy scales with resistance level *in vitro*

We first analysed 51 *H. pylori* isolates recovered from our real-world cohort (Table 1), ensuring the resistance profile reflected clinical reality rather than selective culture bias (Table S4). Among these unselected isolates, 62.7% (32/51) met EUCAST criteria for CLA resistance (MIC of CLA > 0.25 µg/mL). Baseline MICs of CLA showed a clear bimodal distribution, segregating isolates into distinct susceptible (MIC of CLA ≤ 0.25 µg/mL, n = 19) and resistant (MIC of CLA ≥ 4 µg/mL, n = 32) populations with no intermediate phenotypes (Figure 1b).

Checkerboard assays revealed synergistic interactions between CBS and CLA (defined by FICI ≤ 0.5) were exclusive to CLA-resistant isolates. Within this resistant group, baseline MIC of CLA correlated inversely with FICI (*r* = −0.714, *p* < 0.0001, *R*^2^ = 0.509), whereas the association among susceptible isolates was weaker (*r* = −0.566, *p* = 0.011, *R*^2^ = 0.321; Figure 1b, Figures S2–S4).

Laboratory-derived strains with stepwise increases in MIC of CLA (ATCC 43504: 0.015625, CLA-R: 16, CLA-32: 512 µg/mL) showed progressively lower FICI values, confirming increasing CBS–CLA synergy with higher baseline resistance (Figure 1c). 23S rRNA sequencing identified the A2143G mutation in 87.5% (28/32) of resistant isolates, A2142G in 3.1% (1/32), and no target mutation in 9.4% (3/32); no association was found between mutation type and resistance magnitude (Figure 1d).

ROC curve analysis (area under the curve, AUC = 0.991) identified an MIC of CLA cut-off of 12 µg/mL as optimal for predicting strong CBS–CLA synergy. Given routine clinical testing uses binary two-fold dilutions, a threshold of 16 µg/mL was selected for subsequent analyses, with sensitivity of 96.4% and specificity of 91.3% (Figure 1e, Table S5). Time-kill assays were performed using two distinct strain panels: the reference strain panel with stepwise graded CLA resistance (0.015625, 16, 512 μg/mL) and the panel of clinical isolates ( 0.0625, 32, 128 μg/mL). Across both groups, the CBS–CLA combination consistently reduced viable counts by > 2 Log_10_CFU/mL, whereas single-agent treatment showed minimal bactericidal activity (Figure S5).

### 3. Prospective validation of the MIC of CLA threshold in an independent clinical cohort

To prospectively validate the clinical relevance of the MIC of CLA threshold identified *in vitro*, we analyzed treatment outcomes in 49 pediatric patients who participated in the liquid checkerboard experiments described in the previous section and had complete follow-up (two patients were lost to follow-up; Table 2). Strikingly, the MIC of CLA threshold of ≥ 16 µg/mL robustly stratified clinical outcomes between BQT and TT. In high-level resistant infections (MIC of CLA ≥ 16 µg/mL), BQT achieved eradication in 96.0% (24/25) of patients, whereas TT failed in all 3 patients (0%), resulting in an absolute risk difference of 96.0% (95% CI, 73.8% to 100%; *p* < 0.001) (Figure 1f, Table S4). In contrast, for CLA-susceptible infections (MIC of CLA ≤ 0.25 µg/mL), eradication rates were high and not significantly different between BQT (100%, 13/13) and TT (80.0%, 4/5; *p* = 0.172). These results demonstrate that the *in vitro*-derived MIC of CLA threshold (MIC of CLA ≥ 16 µg/mL) accurately identifies patients who will benefit most from bismuth-based therapy.

To further assess the robustness of this MIC threshold, we performed bootstrap resampling (1,000 iterations) on the checkerboard dataset from which the threshold was derived. The median AUC for predicting CBS–CLA synergy remained 0.98 (95% bootstrap *CI*: 0.96–1.00), with sensitivity 94% (95% *CI*: 86–99%) and specificity 91% (95% *CI*: 82–97%), confirming that the MIC ≥ 16 μg/mL threshold is stable and not driven by outlier observations (Figure S6a, b). Additionally, to address potential selection bias in the clinical validation cohort, we performed 1:1 propensity score matching using age, sex, and 23S rRNA mutation status as covariates. Following matching, all covariates achieved adequate balance (standardized mean differences < 0.1), and the superiority of BQT over TT in patients with MIC ≥ 16 μg/mL remained robust (Figure S6c). These complementary analyses collectively support the translational utility of the MIC of CLA ≥ 16 μg/mL threshold for guiding bismuth-based therapy.

### 4. Iron metabolism and energy states differ between CLA-resistant and susceptible strains

To unbiasedly identify the primary biological processes affected by CBS in CLA-resistant *H. pylori*, we first performed transcriptomic profiling of the high-level resistant strain CLA-32 (MIC of CLA = 512 µg/mL) following CBS exposure. Functional enrichment analysis of differentially expressed genes revealed a strong and coherent over-representation of iron-related processes. GO analysis highlighted significant enrichment in iron ion transport, metal ion homeostasis, and redox-associated metabolic functions, while KEGG pathway analysis identified iron acquisition and energy metabolism pathways as dominant functional categories (Figure S7).

Guided by this enrichment signature, differential expression analysis further revealed coordinated repression of the global iron regulator *fur* and multiple iron-dependent metabolic genes (*feoA*, *frdB*, *fbcF*, *frpB*), alongside compensatory upregulation of alternative iron-uptake genes (*feoB*, *fecA*) (Figure 2a). Notably, the discordant regulation of the canonical Feo system—characterised by *feoA* repression but concomitant *feoB* upregulation—argues against a simple iron-starvation response and instead indicates CBS induces a state of iron dyshomeostasis, in which iron sensing, uptake, and utilisation become uncoupled. Together, these transcriptomic changes point to a profound disruption of Fur-governed iron homeostasis rather than a simple iron-replete or iron-starved state. Consistent with these transcriptional signals, both laboratory-derived and clinical CLA-resistant strains exhibited significantly higher intracellular iron content than susceptible strains (Figure 2b). Basal expression of *fur* was markedly elevated in CLA-resistant reference strains and clinical isolates (Figure 2c), indicating an iron-dependent adaptive state associated with CLA resistance.

**Figure 2.**
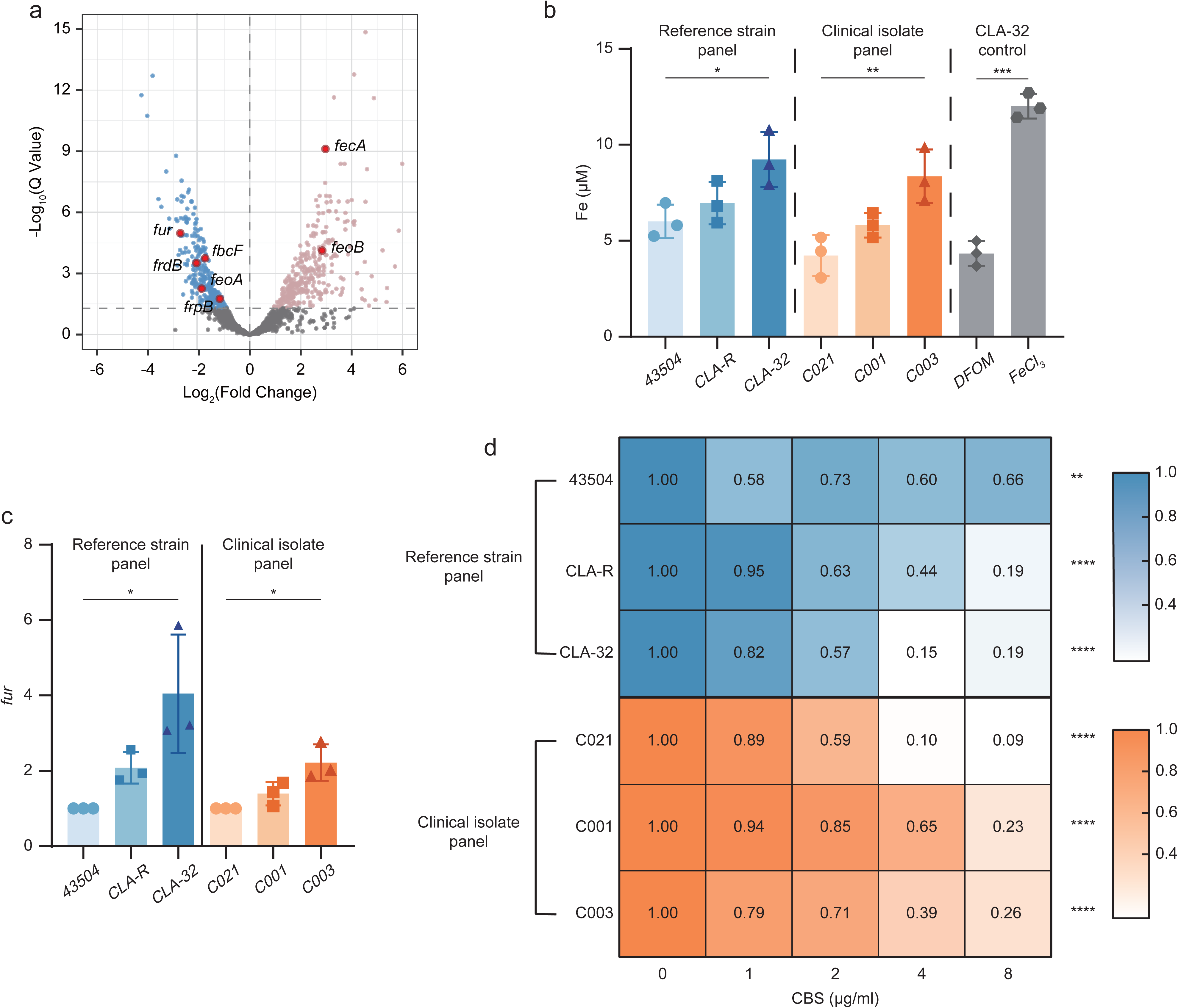
Colloidal bismuth subcitrate (CBS) disrupts Fur-dependent iron homeostasis in clarithromycin (CLA)-resistant *H. pylori*. **(a)** Volcano plot showing differentially expressed genes in the high-level clarithromycin (CLA)-resistant strain CLA-32 following overnight exposure to CBS (8 μg/mL). Red and blue dots indicate significantly up- and down-regulated genes, respectively (|Log2 (fold change)| ≥ 1, adjusted Q value < 0.05). Selected genes involved in iron homeostasis and Fur regulation are highlighted. **(b)** Total cellular iron content measured in reference strains, clinical isolates, and the CLA-32 strain under indicated conditions. Deferoxamine mesylate (DFOM) and ferric chloride (FeCl_3_) were used as iron-depleted and iron-replete controls, respectively. Data are shown as mean ± SD. Statistical significance was assessed using one-way ANOVA with post hoc multiple-comparison correction (\**p* < 0.05; \*\**p* < 0.01; \*\*\**p* < 0.001). **(c)** Basal expression levels of *fur* in reference strains and clinical isolates determined by RT-qPCR. Expression levels were normalized to *gyrB*. Data represent mean ± SD. **(d)** Dose-dependent effects of CBS on *fur* expression across reference strains and clinical isolates. Heatmap values represent relative *fur* expression normalized to the untreated condition (CBS = 0 μg/mL) for each strain. Statistical significance of CBS-dependent repression was evaluated using one-way ANOVA (\*\**p* < 0.01; \*\*\*\**p* < 0.0001).

CBS-mediated repression of *fur* was further validated by RT-qPCR, demonstrating a dose-dependent effect across both reference strains and clinical isolates (Figure 2d). Functionally, CBS treatment significantly reduced intracellular LIPs—a parameter we quantified here for the first time in *H. pylori* using calcein-AM fluorescence. This effect closely paralleled that of the iron chelator DFOM, validating the biological relevance of our LIP measurements (Figures 3a, 3c). CBS also induced a pronounced dose-dependent reduction in intracellular ATP levels, with a 63.2 ± 10.5% decrease observed within 4 hours in CLA-resistant strains (Figures 3b). These findings indicate CBS disrupts Fur-dependent iron homeostasis and impairs energy metabolism in CLA-resistant *H. pylori*.

**Figure 3.**
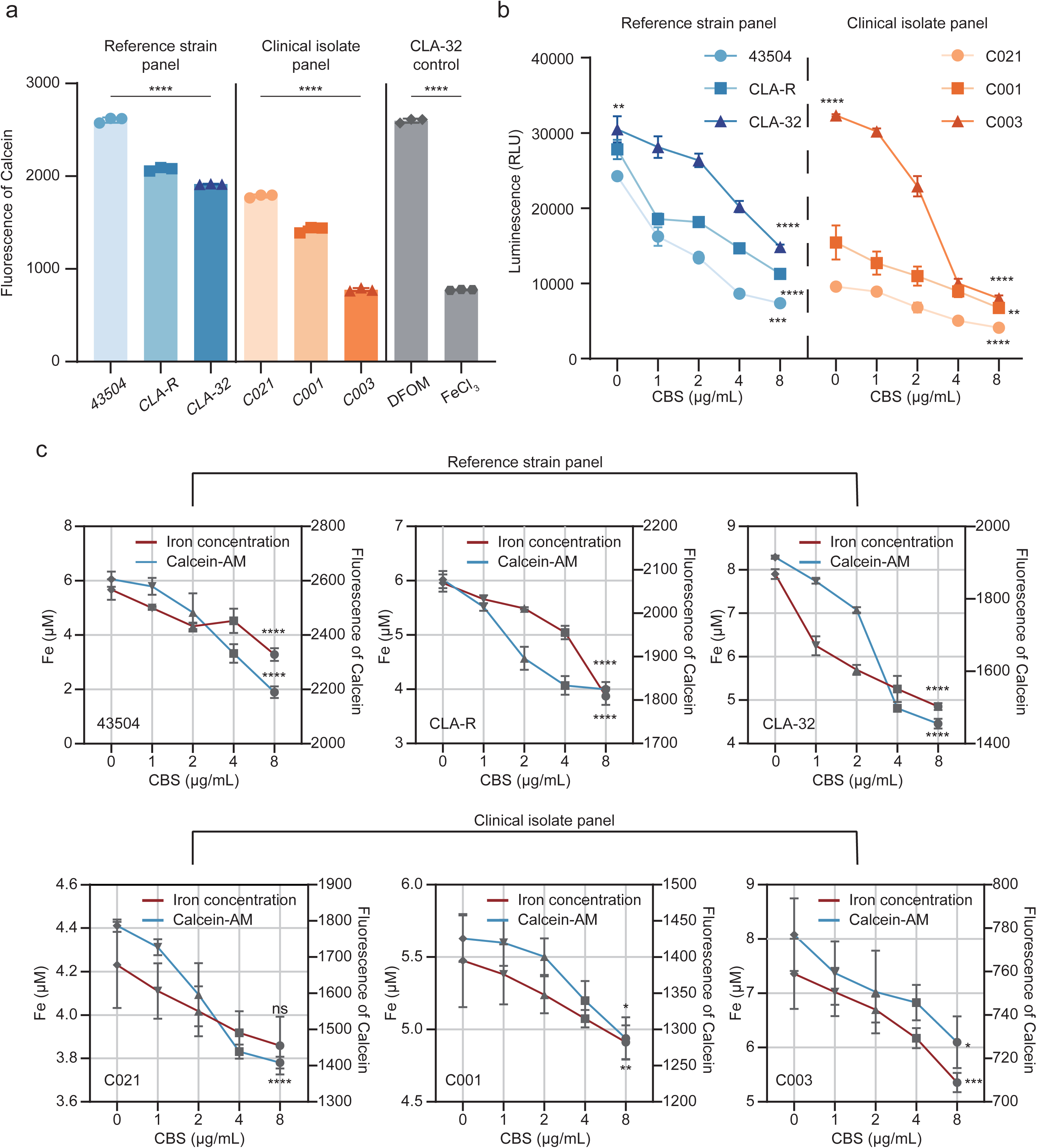
Colloidal bismuth subcitrate (CBS) induces depletion of labile iron pools (LIP) and impairs energy metabolism in clarithromycin (CLA)-resistant *H. pylori*. **(a)** Intracellular LIP assessed using the calcein-AM fluorescence probe in reference strains, CLA-resistant derivatives, and clinical isolates. Reduced fluorescence indicates depletion of bioavailable intracellular iron. Deferoxamine mesylate (DFOM) and ferric chloride (FeCl_3_) were included as iron-depleted and iron-replete controls, respectively. Data are shown as mean ± SD. **(b)** Dose-dependent reduction of intracellular ATP levels following CBS exposure in reference strains and clinical isolates, measured by luminescence-based assays. ATP levels are expressed as relative luminescence units (RLU). Data represent mean ± SD. **(c)** Concurrent changes in total intracellular iron concentration and calcein-AM fluorescence in response to increasing concentrations of CBS. Iron concentrations were quantified following bacterial lysis, while calcein-AM fluorescence reflects labile iron availability. Both parameters decline with CBS treatment, consistent with impaired iron utilisation accompanied by disruption of cellular energy metabolism rather than simple iron chelation. Data are shown as mean ± SD. Statistical significance was assessed using one-way ANOVA. ns, not significant; \**p* < 0.05; \*\**p* < 0.01; \*\*\**p* < 0.001; \*\*\*\**p* < 0.0001.

To directly assess whether iron availability determines the interaction between CBS and CLA, we manipulated intracellular iron levels pharmacologically. Iron chelation with DFOM resulted in a stepwise reduction of MICs of CLA in CLA-resistant reference strains and clinical isolates, whereas CLA-susceptible strains were primarily growth inhibited at low DFOM concentrations (Figure 4a). Conversely, exogenous iron supplementation with FeCl_3_ completely abolished CBS-mediated reductions in MICs of CLA in both reference strains and clinical isolates, restoring MIC of CLA values to those observed in the absence of CBS (Figure 4b). These findings demonstrate that CBS-associated additivity and synergy with CLA are strictly dependent on iron availability.

**Figure 4.**
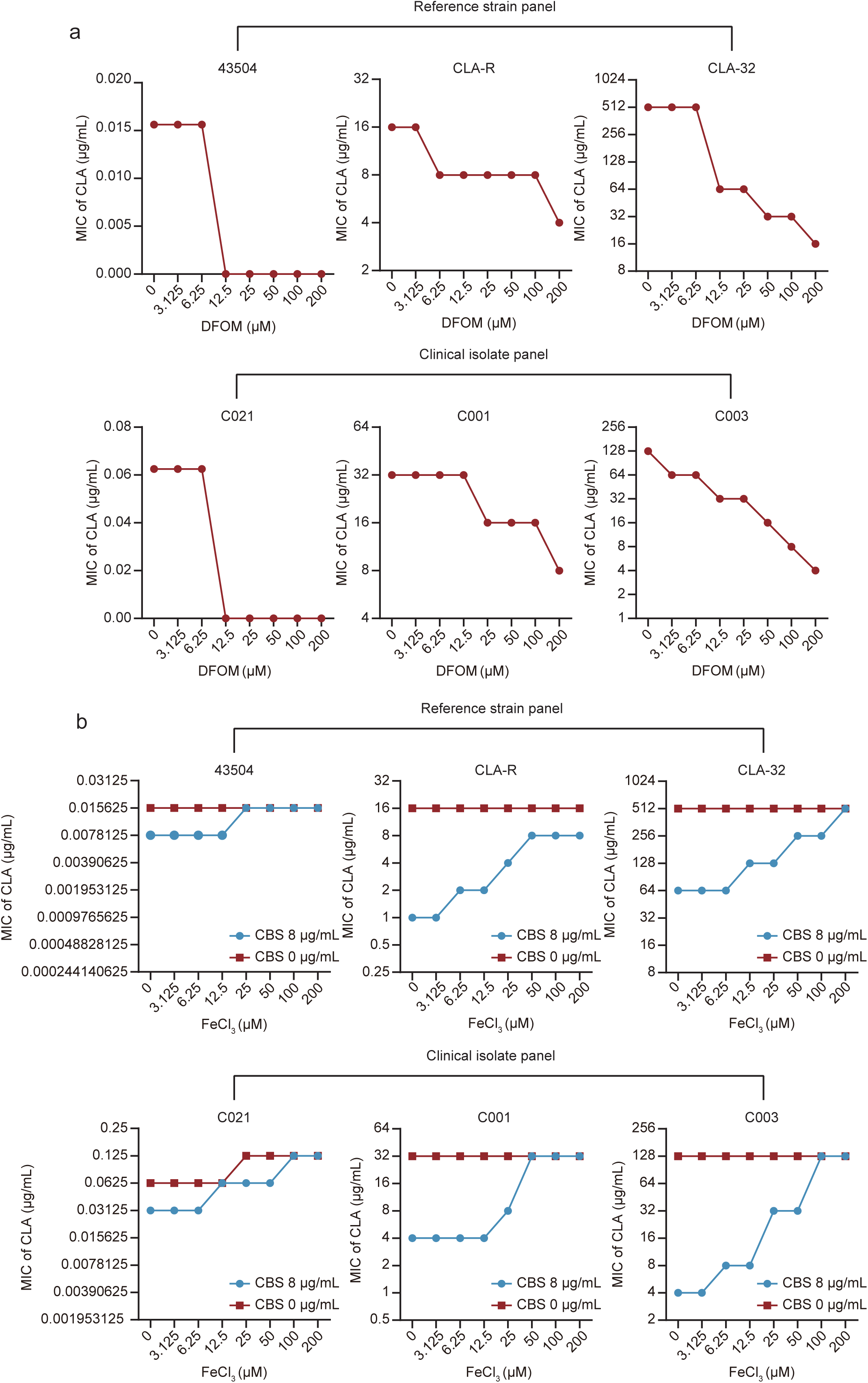
Iron availability determines the interaction between colloidal bismuth subcitrate (CBS) and clarithromycin (CLA). **(a)** Effects of iron chelation on CLA susceptibility. Minimum inhibitory concentrations (MICs) of CLA were determined n reference strains, CLA-resistant derivatives, and clinical isolates in the presence of increasing concentrations of the iron chelator deferoxamine mesylate (DFOM). While low-level iron chelation (12.5 μM DFOM) markedly inhibited the growth of CLA-susceptible strains, progressive iron limitation resulted in a stepwise reduction of MICs of CLA in CLA-resistant strains, indicating iron-dependent sensitization. **(b)** Antagonistic effects of iron supplementation on CBS-mediated additivity and synergy. MICs of CLA were determined in the absence or presence of CBS (8 μg/mL) under increasing concentrations of ferric chloride (FeCl_3_). In both reference strains and clinical isolates, exogenous iron supplementation completely abolished the CBS-induced reduction in MICs of CLA, restoring MIC values to levels observed without CBS. These results indicate that CBS-mediated additivity and synergy with CLA are strictly dependent on iron availability. Data are representative of at least three independent experiments.

### 5. Enhanced efflux activity in CLA-resistant strains is not directly modulated by bismuth

To characterise efflux activity in *H. pylori*, we employed a continuous real-time monitoring approach, representing the first application of dynamic EtBr accumulation and efflux assays in this pathogen. CLA-resistant isolates showed stepwise increases in EtBr efflux capacity that correlated positively with baseline MIC of CLA (Figure 5a–d). Notably, EtBr accumulation reached a plateau at ∼20 minutes in the reference strain-derived panel, whereas it continued to rise throughout observation in clinical CLA-resistant isolates (Figure 5a–d).

**Figure 5.**
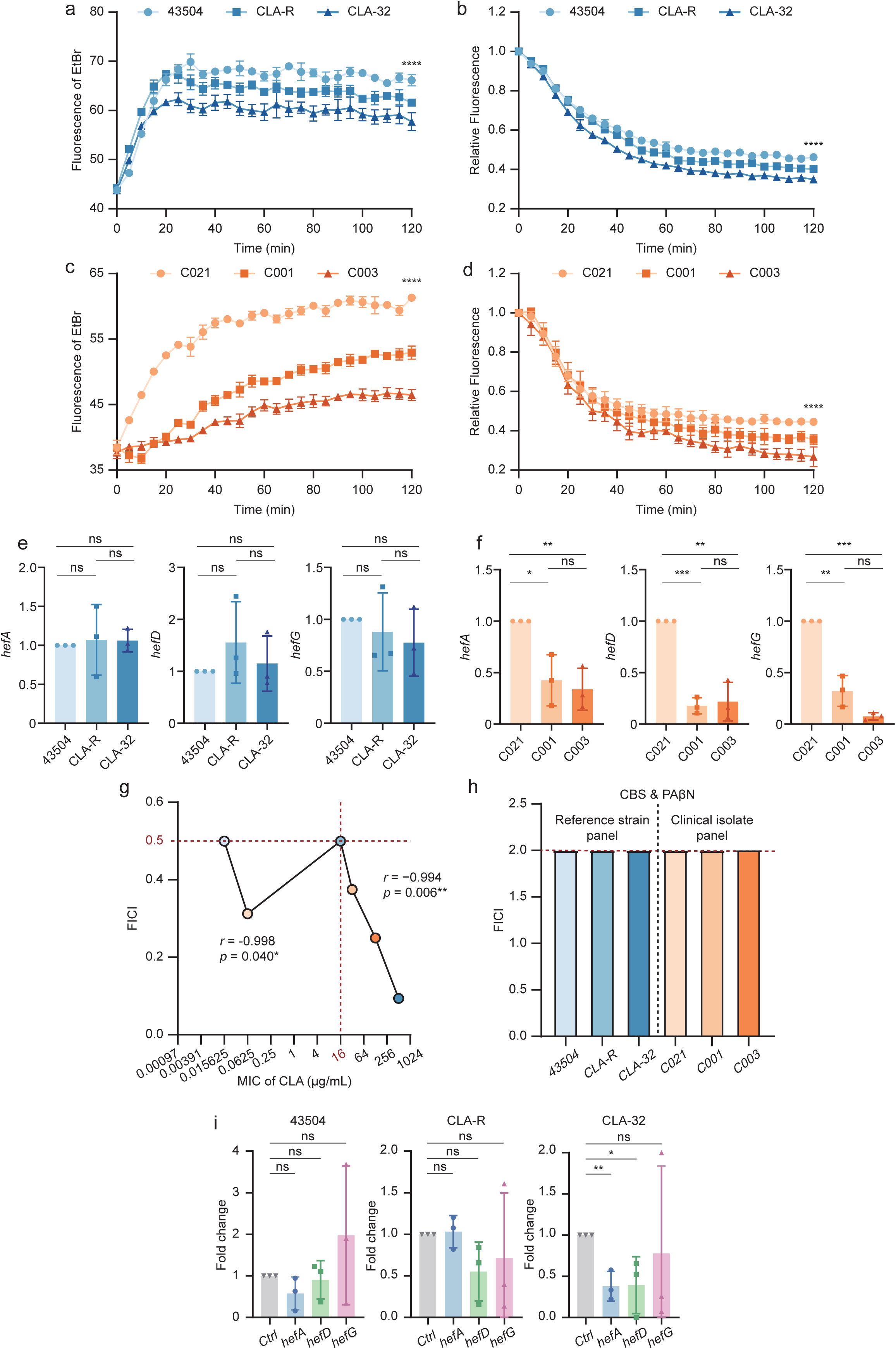
Colloidal bismuth subcitrate (CBS)–clarithromycin (CLA) synergy is independent of RND efflux pump activity in *H. pylori*. **(a–d)** Ethidium bromide (EtBr) accumulation **(a, c)** and efflux **(b, d)** assays in reference strains and clinical isolates. Fluorescence intensity reflects intracellular EtBr levels and serves as an inverse proxy for global efflux capacity. CLA-resistant strains exhibited reduced EtBr accumulation and enhanced EtBr efflux, consistent with increased efflux activity. **(e, f)** Expression levels of RND efflux pump components (*hefA*, *hefD*, and *hefG*) in reference strains **(e)** and clinical isolates **(f)**, determined by RT-qPCR. In clinical isolates, expression of all three genes was significantly reduced with increasing CLA resistance. Data are presented as mean ± SD. **(g)** Fractional inhibitory concentration indices (FICI) for the combination of the efflux pump inhibitor phenyl-arginine-β-naphthylamide (PAβN) and CLA in reference-derived and clinical isolate panels, with strains ordered by increasing MIC of CLA. PAβN exhibited synergistic interactions with CLA regardless of resistance status, and FICI values showed a significant negative correlation with MIC of CLA when MIC of CLA ≥ 16 μg/mL, indicating enhanced synergy in high-level resistant strains. **(h)** FICI values for the combination of PAβN and CBS in reference strains and clinical isolates. All strains displayed indifferent interactions (FICI = 2), indicating that CBS activity is not mediated through inhibition of RND efflux pumps. **(i)** mRNA expression of RND efflux pump components in reference strains (43504, CLA-R, CLA-32) following treatment with the efflux pump inhibitor PAβN PAβN reduced *hefA* and *hefD* expression specifically in the high-level resistant strain CLA-32. Statistical significance was assessed using one-way ANOVA, Student’s t-test, and Pearson’s linear regression, as appropriate; ns, not significant; \**p* < 0.05; \*\**p* < 0.01; \*\*\**p* < 0.001; \*\*\*\**p* < 0.0001.

This enhanced efflux phenotype occurred without transcriptional upregulation of major RND efflux pump genes (*hefA*, *hefD*, *hefG*); in fact, expression of all three genes decreased significantly in clinical isolates as CLA resistance increased (Figure 5e, f). Checkerboard assays showed the RND efflux inhibitor phenyl-arginine-β-naphthylamide (PAβN) had synergistic activity with CLA in both susceptible and resistant strains. For isolates with MIC of CLA ≥ 16 µg/mL, there was a strong inverse correlation between FICI and baseline MIC (*r* = −0.994, *p* = 0.006), indicating enhanced synergy in highly resistant strains (Figure 5g). In contrast, PAβN showed an indifferent interaction with CBS (FICI = 2.0, Figure 5h). RT-qPCR confirmed PAβN selectively suppressed *hefA* and *hefD* expression in the high-level resistant strain CLA-32 (Figure 5i). To further assess whether CBS modulates efflux function, we treated high-level resistant strains CLA-32 (laboratory-derived) and C003 (clinical) with increasing CBS concentrations. CBS did not significantly alter EtBr efflux activity in either strain, whereas the protonophore CCCP reduced efflux by ∼42.3 ± 4.2% (Figure 6a–d, S8), confirming CBS does not directly inhibit proton motive force (PMF)-dependent efflux. Transcriptional profiling revealed strain-specific regulatory effects of CBS on RND efflux system components, rather than global inhibition. In the laboratory-derived high-level resistant strain CLA-32, CBS treatment selectively suppressed *hefG* expression but had no significant impact on *hefA* or *hefD*. Conversely, in the clinical high-level resistant isolate C003, CBS downregulated *hefD* and *hefG* while actually upregulating *hefA* (Figure 6e). This divergent pattern confirms that CBS does not exert uniform inhibitory effects on major RND efflux pumps, particularly failing to suppress the primary pump *hefA* in either strain. These data demonstrate CBS-mediated CLA potentiation is independent of direct RND efflux pump inhibition.

**Figure 6.**
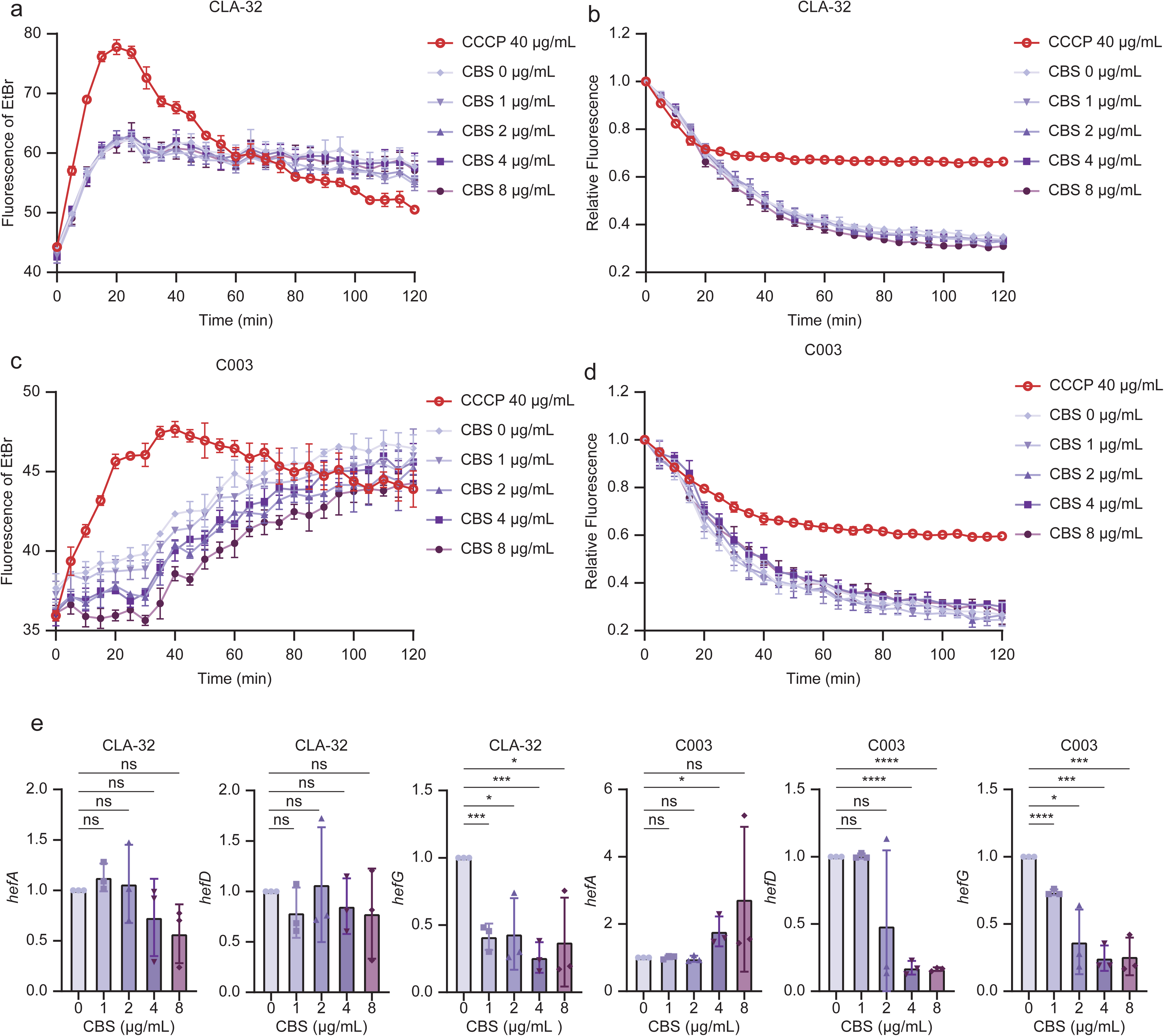
Efflux pump function and expression profiling in high-level clarithromycin (CLA)-resistant strains **(a–d)** Ethidium bromide (EtBr) accumulation **(a, c)** and active efflux kinetics **(b, d)** in the highest-level clarithromycin-resistant strains CLA-32 (reference panel) and C003 (clinical isolate panel) treated with increasing concentrations of CBS (0 to 8 μg/mL). Carbonyl cyanide m-chlorophenyl hydrazone (CCCP, 40 μg/mL) was used as a positive control for efflux inhibition. CBS treatment did not significantly alter efflux function in either strain, while CCCP only partially inhibited efflux. **(e)** mRNA expression of RND efflux pump components (*hefA*, *hefD*, and *hefG*) in CLA-32 and C003 following CBS treatment, determined by RT-qPCR. CBS showed strain-specific effects: in CLA-32, only *hefG* expression was suppressed, while in C003, *hefD* and *hefG* were downregulated but *hefA* was upregulated. Data are presented as mean ± SD relative to untreated controls (0 μg/mL CBS). Statistical significance was assessed using Student’s t-test; ns, not significant; \**p* < 0.05; \*\*\**p* < 0.001; \*\*\*\**p* < 0.0001.

### 6. CBS-associated synergy is specific to CLA resistance and reflects a resistance-linked physiological state

To test whether CBS potentiation generalises beyond CLA resistance, we generated isogenic one-step-resistant derivatives of ATCC 43504 targeting AMO, LEV, RIF, and TET, alongside our established CLA-resistant (CLA-R) strain. One-step selection recapitulates clinical high-level resistance without confounding secondary mutations; intrinsic MET resistance in ATCC□43504 precluded generating MET-resistant derivatives.

Checkerboard assays confirmed CBS synergized exclusively with CLA in CLA-R strains, showing no consistent potentiation in AMO, LEV, MET, RIF, or TET-resistant backgrounds. The CLA-R strain exhibited a non-linear, threshold-dependent synergy profile matching clinical isolates, while other resistant derivatives showed indifferent or antagonistic interactions with CBS (Figure 7a; Table□S6). This specificity indicates CBS synergy is tied to the unique physiological signature of CLA resistance, not a general stress response.

**Figure 7.**
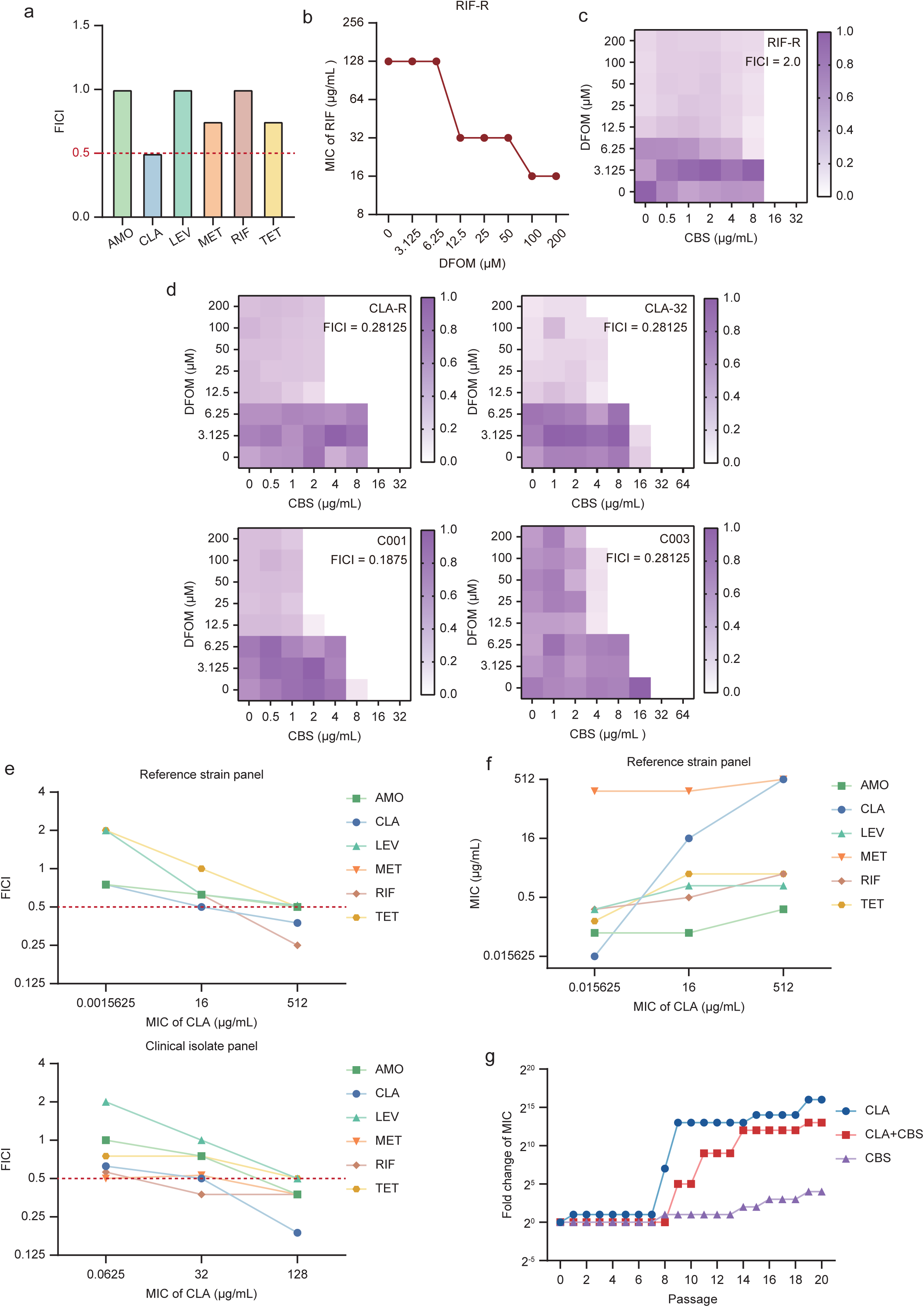
CBS-associated synergy is specific to clarithromycin (CLA) resistance and reflects a resistance-associated physiological state **(a)** Fractional inhibitory concentration indices (FICI) for the combination of colloidal bismuth subcitrate (CBS) with different antibiotics in isogenic derivatives of *H. pylori* ATCC□43,504 generated by one-step selection for resistance to amoxicillin (AMO), CLA, levofloxacin (LEV), metronidazole (MET), rifampicin (RIF), or tetracycline (TET). Synergistic interaction (FICI□≤□0.5, dashed line) was observed exclusively in the CLA-resistant derivative. **(b)** Effect of iron chelation on RIF resistance. Increasing concentrations of deferoxamine mesylate (DFOM) resulted in a stepwise reduction of MIC of RIF in RIF-resistant (RIF-R) strain. **(c)** Checkerboard analysis of CBS and DFOM in RIF-R strain. Heatmap representation of growth inhibition with corresponding FICI values indicating indifferent interaction (FICI□=□2.0). **(d)** Checkerboard analyses of CBS and DFOM in CLA-resistant strains from both reference-derived (CLA-R, CLA-32) and clinical isolate panels (C001, C003). All CLA-resistant strains exhibited synergistic interactions (FICI□≤ □0.5) with characteristic right-angled (L-shaped) isoboles, consistent with partial mechanistic overlap along an iron-dependent axis. **(e)** Relationship between CBS-associated FICI values and baseline MIC of CLA in reference strain (top) and clinical isolate (bottom) panels. Synergy was selectively enhanced with increasing levels of CLA resistance. **(f)** Corresponding MIC values of multiple antibiotics measured across stepwise CLA–resistant derivatives of the reference strain ATCC□43504, plotted against the MIC of CLA, illustrating the progressive increase in resistance to other antibiotics during the induction of increasing levels of CLA resistance. **(g)** Stepwise selection experiments showing fold changes in MICs over serial passages: MICs of CLA were measured for cultures passaged with CLA alone and the CBS+CLA combination, while MICs of CBS were measured for cultures passaged with CBS alone. CBS did not prevent the emergence of CLA resistance but delayed the development of high-level CLA resistance (MIC ≥ 16 μg/mL) by five passages. Serial passage with CBS alone led to reduced susceptibility to CBS, but this did not affect its ability to delay CLA resistance evolution. Data are representative of at least three independent experiments. Where applicable, values are presented as mean □±□SD.

Iron chelation with DFOM reduced MICs of CLA in CLA-R strain and RIF in RIF-R strain, but had no consistent effect on other resistant backgrounds (Figure 7b; Figure S9). Checkerboard analysis demonstrated that CBS combined with RIF exhibited an indifferent interaction in the RIF-R strain (FICI = 2.0). In contrast, the CBS–DFOM combination displayed synergistic interactions (FICI ≤ 0.5) exclusively in CLA-resistant strains, with characteristic right-angled inhibition patterns, supporting a mechanistic interaction distinct from simple iron limitation (Figure 7c, d).

Across reference and clinical strains, CBS synergy correlated inversely with baseline MIC of CLA (Figure 7e), while CLA resistance induction in ATCC□43504 conferred cross-resistance to other antibiotics (Figure 7f). These findings indicate bismuth’s resistance-suppressive effects extend beyond CLA to other classes only in CLA-resistant backgrounds.

Iron availability directly modulated this expanded synergistic phenotype: DFOM reduced multiple antibiotic MICs in CLA-resistant isolate C003, while exogenous iron abolished CBS-mediated potentiation (Figure□S9). Together, these data demonstrate iron availability negatively regulates CBS’s capacity to potentiate antibiotic activity in CLA-resistant *H. pylori*.

### 7. Bismuth delays emergence of high-level CLA resistance

Serial passage of CLA-susceptible *H. pylori* ATCC 43504 under CLA selective pressure resulted in discontinuous, large-step increases in MIC of CLA (0.03125 to 2 to 128 µg/mL), indicative of stepwise acquisition of high-level resistance (Figure 7g). Co-exposure to CBS delayed the emergence of high-level CLA resistance (MIC ≥ 16 µg/mL) by five serial passages, although resistance ultimately developed. Gradual increases in MIC of CBS were observed during co-passaging, consistent with adaptive changes in bismuth susceptibility, but this did not abrogate the delay in high-level CLA resistance emergence (Figure 7g).

## Discussion

The sustained efficacy of BQT against CLA-resistant *H. pylori* has long been recognized but mechanistically unexplained.^11–13, 16^ Our study resolves this paradox by demonstrating that high-level CLA resistance (MIC ≥ 16 μg/mL) defines a metabolically distinct, iron-dependent physiological state that creates a targetable vulnerability uniquely exploitable by bismuth.^15, 28^ Mechanistically, CLA resistance upregulated the iron regulator *fur*, increasing intracellular iron and ATP—creating dependence on iron homeostasis. Bismuth disrupts this finely balanced state by inducing iron dyshomeostasis rather than simple chelation: CBS treatment reduced total intracellular iron while paradoxically increasing LIPs, indicating misallocation of iron stores rather than uniform depletion. This mechanistic distinction was further supported by the L-shaped synergistic interaction between CBS and the iron chelator DFOM—a pattern indicative of partial mechanistic overlap along an iron-dependent axis rather than identical pathways. The resulting profound ATP depletion selectively compromises resistant bacteria whose survival depends on elevated iron and energy flux to support resistance mechanisms such as efflux.^15, 24, 29^ This dual metabolic injury was specific to CLA resistance, as resistance to other antibiotic classes did not confer bismuth synergy.

These effects were context-dependent and species-specific. Unlike *Pseudomonas aeruginosa*, in which bismuth directly inhibits proton motive force–dependent efflux pumps, Helicobacter *pylori* possesses a heterogeneous efflux repertoire comprising both ATP-dependent and PMF-driven systems.^26, 30, 31^ Our findings suggest that bismuth-induced energy depletion destabilizes energy-intensive resistance architectures rather than fully restoring antibiotic susceptibility, thereby facilitating effective combination therapy.

Clinically, this translates into a simple, actionable threshold. In the largest real-world cohort of CLA-resistant paediatric infections (n = 4,610; 1,844 with follow-up), BQT achieved 93.1% eradication versus 68.8% with TT (*p* = 0.017). Bismuth–CLA synergy scaled precisely with resistance level, yielding an optimal MIC of CLA cutoff of ≥ 16 μg/mL (AUC = 0.991). Prospectively validated in an independent cohort of 49 patients, this threshold robustly stratified outcomes: BQT cured 96% of high-level resistant patients (MIC ≥16 μg/mL) versus 0% with triple therapy (*p* < 0.001). Notably, a recent study using Etest identified a 12 μg/mL threshold for predicting BQT efficacy,^28^ closely matching our ROC-derived optimum of 12 μg/mL. Given routine clinical use of two-fold dilutions, we selected 16 μg/mL as a pragmatic cut-off—validated here by its strong association with curative efficacy.

With ethical constraints now precluding placebo-controlled randomized trials in patients with proven high-level resistance, the next logical step is a multi-center, all-age, biomarker-stratified prospective cohort study to further validate and generalize these findings. Given the large effect size (relative risk reduction >5-fold) and clear dose–response gradient, our findings will satisfy GRADE criteria for upgrading observational evidence to high quality (Grade 1++).^32^

Our work builds upon and significantly extends previous observations of a bimodal MIC distribution in CLA-resistant *H. pylori*.^28, 33^ While prior studies attributed this bimodality to macrolide exposure history, we propose—for the first time—that this high-level resistance cluster represents a fundamental transition in bacterial physiology. This conceptual shift from epidemiological association to biological state change provides a novel framework for understanding treatment responses and moves beyond binary resistance classification toward a quantitative, vulnerability-oriented paradigm.

Importantly, the principle demonstrated here is likely not unique to CLA. Each antibiotic resistance mechanism probably imposes its own distinctive metabolic burden. Our focus on CLA, driven by its high prevalence, should not obscure the broader reality: resistance is heterogeneous, and the vulnerabilities it creates are likely just as diverse.^25^ The future of H. pylori eradication may lie in mapping these resistance-specific metabolic dependencies and developing a toolkit of targeted adjuncts—of which bismuth is simply the first, but by no means the only, exemplar. Our demonstration that the iron chelator deferoxamine also targets high-level CLA-resistant strains (and, notably, RIF-resistant strains) further validates iron dependence as a core vulnerability and hints at broader applicability.

Several limitations warrant consideration. The prospective validation cohort was modest in size, and the predictive performance of the MIC ≥ 16 μg/mL threshold requires confirmation in larger, multi-center studies. The genetic intractability of *H. pylori* precluded definitive assignment of single-gene causality.^34^ Nevertheless, the strong concordance across clinical, phenotypic, transcriptomic, and evolutionary datasets provides compelling support for the proposed model.

These limitations do not diminish the immediate translational implications. Rather than applying bismuth empirically to all resistant infections, clinicians can now use a quantitative MIC to identify patients who benefit most—a strategy aligned with antimicrobial stewardship. We propose a pragmatic MIC-guided framework (Figure 8): for MIC ≥16 μg/mL, BQT should be strongly preferred as first-line therapy (96% efficacy vs 0% with TT); for intermediate MIC (0.5–8 μg/mL), bismuth-based regimens remain a rational option.

**Figure 8.**
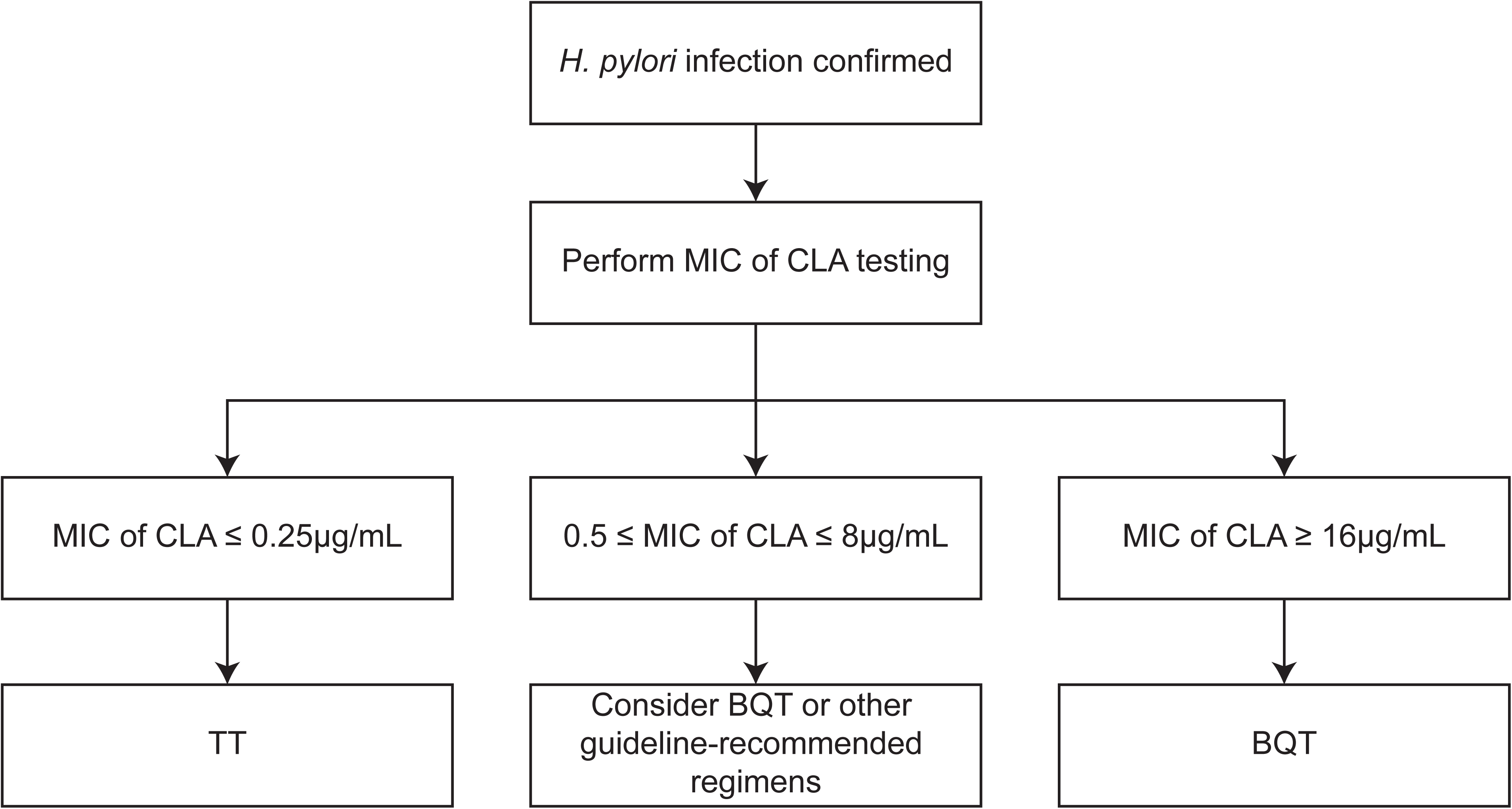
Quantitative clarithromycin (CLA) susceptibility testing is performed following confirmation of *H. pylori* infection. MIC of CLA ≤ 0.25 µg/mL (Susceptible): Standard triple therapy (TT) remains a highly effective and appropriate first-line option. MIC of CLA ≥ 16 µg/mL (High-level resistant): Bismuth-containing quadruple therapy (BQT) should be strongly prioritized as first-line treatment. In our clinical validation cohort, BQT achieved 96.0% eradication in this group, whereas TT failed in all cases (0%). MIC of CLA 0.5–8 µg/mL (Intermediate resistant): Both BQT and TT showed 100% eradication in our limited cohort; however, BQT represents a prudent, guideline-endorsed choice for resistant infections. This stratified approach aims to optimize outcomes by matching the intensity of therapy to the level of resistance, moving beyond binary susceptibility categorization toward precision stewardship.

In summary, our study reframes antibiotic resistance not as an endpoint but as a therapeutic opportunity. By demonstrating that the metabolic cost of resistance can be therapeutically exploited, we provide a blueprint for mechanism-informed precision therapy and lay the foundation for a new era of resistance-targeted eradication.

## Data availability statement

Experimental data are available from the corresponding author (Ying Huang) upon reasonable request. The CLA-32 transcriptome data have been deposited in the NCBI Sequence Read Archive (accession number: PRJNA1392096).

## Ethics statements

## Patient consent for publication

Not applicable.

## Ethics approval

The study was approved by the Institutional Ethics Committee of Children’s Hospital of Fudan University (approval No. [2017]186) and was conducted in accordance with the Declaration of Helsinki. Written informed consent was obtained from the guardians of all participants.

## Author contributions

Chunmeng He: Conceived and designed the entire study, performed all experimental work, analyzed and interpreted all data, drafted the full manuscript, and responded to reviewer comments.

Ying Huang: Secured funding and resources, provided overall study supervision, and offered critical intellectual input on study design, including guidance on the inclusion of deferoxamine mesylate (DFOM) as a reverse validation tool.

All authors: Reviewed, edited, and approved the final version of the manuscript.

## Acknowledgements

We thank the patients and their guardians for participating in this study, as well as the staff at the Department of Gastroenterology, Children’s Hospital of Fudan University. We also acknowledge Associate Prof. Liangdong Lyu for providing experimental resources. We also thank Dr. Ying Zhou for providing the retrospective data.

