## Supplementary Methods for "High-level clarithromycin resistance: a metabolic vulnerability exploited by bismuth in *Helicobacter pylori*"

1. ****Clinical Cohort****

**A single-center, retrospective study was conducted at Children's Hospital of Fudan University between 2019 and 2024. Patients aged 6–18 years with culture-confirmed or histologically/biochemically validated *H. pylori* infection and no prior eradication therapy were included.**^1^ **Endoscopic biopsy specimens for culture were collected selectively based on clinical indication (e.g., persistent symptoms, high suspicion of treatment failure) rather than universally, consistent with routine clinical practice. Eradication regimens, clinical outcomes, and antimicrobial susceptibility data were retrieved from electronic medical records.**

2. ****Bacterial Isolates and Culture****

**Clinical isolates were prospectively collected from May to August 2024 (n = 51). *H. pylori* strains were cultured on Columbia agar supplemented with 5% defibrinated horse blood under microaerophilic conditions (5% O_2_, 10% CO_2_, 85% N_2_) at 37°C for at least 72 hours. Identification was confirmed by Gram staining (curved, Gram-negative rods) and positive results for urease, catalase, and oxidase activities.**

3. ****23S rRNA Mutation Detection****

**Genomic DNA was extracted from *H. pylori* isolates using the QIAamp DNA Mini Kit (Qiagen, Hilden, Germany). The 23S rRNA gene was amplified via PCR using specific primers (Forward: 5’-GAAGAGTTTGATCATGGCTC-3’, Reverse: 5’-CTACGGCTACCTTGTTACGA-3’). PCR products were purified and subjected to Sanger sequencing (Sangon Biotech, Shanghai, China). Sequencing data were aligned with the reference *H. pylori* 26695 sequence (GenBank Accession No.: U27270)** ^2^ **using SnapGene software (version 6.0.2, GSL Biotech, San Diego, USA) to identify point mutations at positions 2142 and 2143.**

4. ****Antimicrobial Susceptibility Testing****

**Minimum inhibitory concentrations (MICs) were determined by broth microdilution according to EUCAST guidelines version 13.0, with incubation at 37°C for 72–96 hours at 120 rpm in a microaerophilic environment. Resistance breakpoints were defined per EUCAST criteria (MIC of CLA ≥ 0.25 μg/mL).**^3^

5. ****Checkerboard Assays****

**In vitro drug interactions were evaluated using checkerboard broth microdilution. The fractional inhibitory concentration index (FICI) was calculated as:**

**FICI = MIC(A/B)/MIC(A) + MIC(B/A)/MIC(B)**

**Interactions were categorized according to EUCAST criteria: synergy (FICI ≤ 0.5), additive effect (0.5 < FICI ≤ 1), and no interaction (1 < FICI ≤ 2). All assays were performed in biological triplicate.**^3^

6. ****Construction of Isogenic Resistant Strains****
   6.1. ****One-Step Selection:****

*H.* ***pylori* ATCC 43504 was subjected to high-concentration antibiotic selection on agar plates to generate clones resistant to amoxicillin (AMO), clarithromycin (CLA), levofloxacin (LEV), tetracycline (TET), and rifampicin (RIF).**^3^

6.2. ****CLA Resistance Gradient:****

**A one-step CLA-resistant progenitor strain was serially passaged in liquid broth with progressively increasing CLA concentrations to generate derivatives with MICs of 16, 64, and 256 μg/mL, consistent with high-level resistance observed in clinical isolates.**

6.3. ****Time-Kill Assays:****

**Bactericidal activity was assessed over 24 hours. A bactericidal effect was defined as a ≥ 2 Log_10_ reduction in colony-forming units (CFU/mL) relative to baseline. All assays were performed in biological triplicate.**^4^

7. ****Metabolic State Quantification****
   7.1. ****Intracellular Iron:****

**Labile iron pools were measured using calcein-AM (20 μM, 30 min loading in HEPES buffer).**^5^ **Total iron content was quantified colorimetrically using the Total Iron Assay Kit (Elabscience, Wuhan, China).**

7.2. ****ATP Content:****

**Intracellular ATP levels were measured using the BacTiter-Glo™ Microbial Cell Viability Assay Kit (Promega, Madison, USA) with an optimized lysis protocol.**

8. ****Efflux Pump Activity Assay****

**Ethidium bromide (EtBr) accumulation and efflux were monitored fluorometrically in HEPES-buffered medium. Carbonyl cyanide m-chlorophenyl hydrazone (CCCP, 40 μg/mL) was used as a positive control for efflux pump inhibition.**^6^

9. ****Transcriptomic Analysis and RT-qPCR****

**Total RNA was extracted using RNAiso Plus (Takara Bio, Otsu, Japan), reverse transcribed to cDNA with the TransScript Uni All-in-One Kit (TransGen Biotech, Beijing, China), and analyzed by RT-qPCR using a QuantStudio 5 system (Thermo Fisher Scientific, Waltham, USA). The *gyrB* gene served as the reference gene, with primer sequences listed in the following table.**^7^ **Relative gene expression was calculated using the 2^⁻ΔΔCT^ method.**

**Primer sequences used for RT-qPCR and mutation detection**

| Gene |  | Primer (5’-3’) |
| --- | --- | --- |
| *gyrB* | F | ATTGGCAAAACCAAAAGTGC |
| *gyrB* | R | CCGTCGTTAAGATACGCCAT |
| *fur* | F | ATTGGCAAAACCAAAAGTGC |
| *fur* | R | CCGTCGTTAAGATACGCCAT |
| *hefA* | F | TATGCCCGCTGTTGA |
| *hefA* | R | GAGGAAATACGACGCTAA |
| *hefD* | F | GGCGTTGATTCTAATGTTTC |
| *hefD* | R | GAAGTGCGTATCCCTTTAC |
| *hefG* | F | CATTTGAGATTGCGTGA |
| *hefG* | R | TCGTTAGCAAGTGGGATA |

10. ****Experimental Evolution****
    10.1. *****H. pylori* ATCC 43504 was serially passaged (20 passages, 48 hours per passage) under three experimental conditions:****
(1) **Medium containing sub-inhibitory clarithromycin (CLA, 1/2 MIC, 0.0156 μg/mL) alone**
(2) **Medium containing sub-inhibitory CLA (1/2 MIC, 0.0156 μg/mL) combined with 4 μg/mL colloidal bismuth subcitrate (CBS)**
(3) **Medium containing sub-inhibitory CBS (1/2 MIC, determined via broth microdilution according to EUCAST guidelines version 13.0) alone**
    10.2. ****MIC monitoring:****
(1) **For conditions 1 and 2, MICs of CLA were measured after each passage to evaluate the rate of resistance emergence**
(2) **For condition 3, MICs of CBS were determined at each passage to monitor adaptive changes in bismuth susceptibility**
11. ****Statistical Analysis****

**Statistical analyses were performed using SPSS 20.0 (IBM, Armonk, USA), GraphPad Prism 9.0 (GraphPad Software, San Diego, USA), and R version 4.5.2 (R Foundation, Vienna, Austria). Normality testing was performed using the Shapiro–Wilk test for sample sizes <500 and Kolmogorov–Smirnov test for sample sizes ≥500. Variances were assessed using Levene's test.Continuous variables were compared using Student’s t-test (two groups) or one-way ANOVA ( ≥ 3 groups). Categorical variables were compared using chi-square tests or Fisher’s exact test, as appropriate. All tests were two-tailed, with *p* < 0.05 considered statistically significant.**

****References****

1. Homan M, Jones NL, Bontems P, et al. Updated joint ESPGHAN/NASPGHAN guidelines for management of Helicobacter pylori infection in children and adolescents (2023). J Pediatr Gastroenterol Nutr 2024;79:758–85. doi:10.1002/jpn3.12314

2. Versalovic J, Osato MS, Spakovsky K, et al. Point mutations in the 23S rRNA gene of Helicobacter pylori associated with different levels of clarithromycin resistance. J Antimicrob Chemother 1997;40:283–6. doi:10.1093/jac/40.2.283

3. The European Committee on Antimicrobial Susceptibility Testing.

Breakpoint tables for interpretation of MICs and zone diameters.

Version 13.0. Växjö: EUCAST; 2023.

Available from: https://www.eucast.org

4. Aguilera-Correa JJ, Urruzuno P, Barrio J, et al. Detection of Helicobacter pylori and the genotypes of resistance to clarithromycin and the heterogeneous genotype to this antibiotic in biopsies obtained from symptomatic children. Diagn Microbiol Infect Dis 2017;87:150–3. doi:10.1016/j.diagmicrobio.2016.03.001

5. Page MG, Mueller C, Hofer B, et al. Enhanced Activity of the Siderophore Monosulfactam BAL30072 (BAL) against Acinetobacter in Iron-Limited Conditions. Abstracts of the Interscience Conference on Antimicrobial Agents and Chemotherapy 2010;50.

6. Paixao L, Rodrigues L, Couto I, et al. Fluorometric determination of ethidium bromide efflux kinetics in Escherichia coli. Journal of biological engineering 2009;3:18–. doi:10.1186/1754-1611-3-18

7. Lee SM, Kim N, Kwon YH, et al. rdxA, frxA, and efflux pump in metronidazole-resistant Helicobacter pylori: Their relation to clinical outcomes. J Gastroenterol Hepatol 2018;33:681–8. doi:10.1111/jgh.13906
