## Supplementary material for "High-level clarithromycin resistance: a metabolic vulnerability exploited by bismuth in *Helicobacter pylori*": Figure S1-S9

**Supplementary Figures**

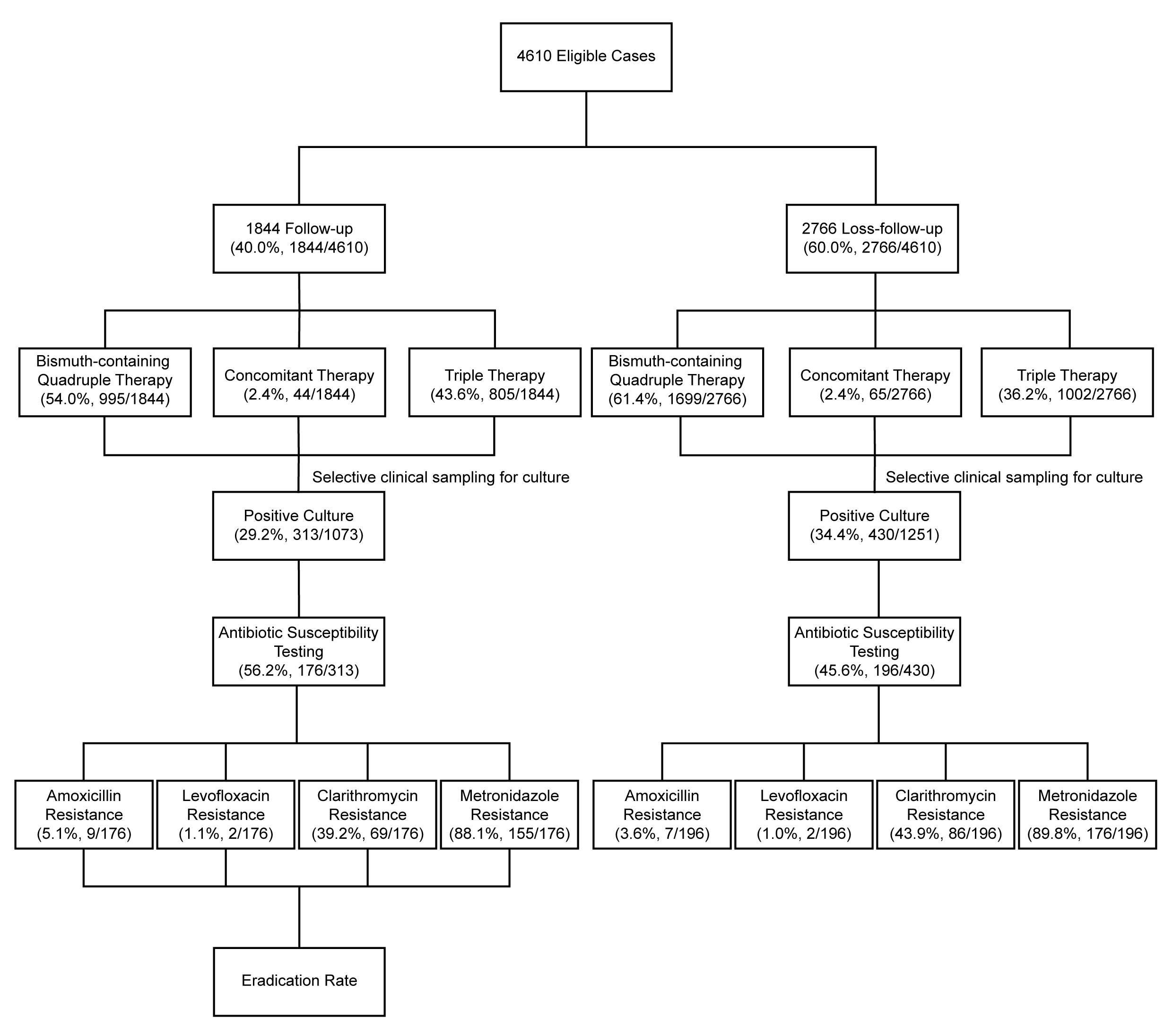

**Figure S1.** Study flow chart illustrating patient selection, treatment regimens, selective culture sampling, antibiotic susceptibility testing, and eradication outcome assessment.

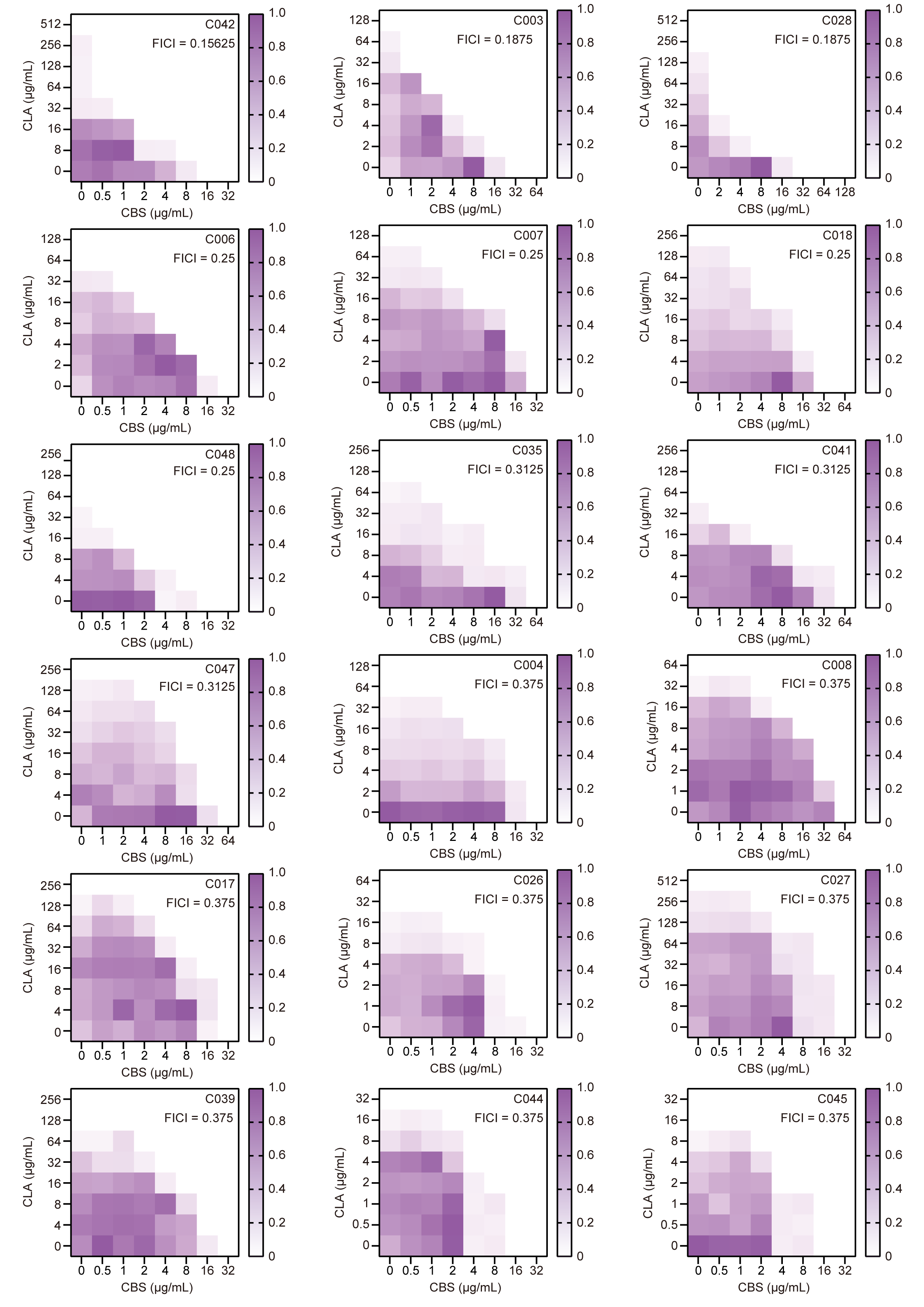

### **Figure S2.** Checkerboard assays showing the interaction between CBS and CLA against selected clinical isolates.

### Checkerboard microdilution assays were performed to evaluate the combined antibacterial activity of CBS and CLA against clinical isolates. Heatmaps show fractional growth inhibition across a range of CBS (x‑axis, μg/mL) and CLA (y‑axis, μg/mL) concentrations. Color intensity indicates the degree of growth inhibition, normalized to the untreated control (white, no inhibition; dark purple, maximal inhibition). Panels shown represent selected isolates exhibiting strong synergy (FICI ≤ 0.375). The corresponding FICI values for each isolate are indicated in the upper right corner of each panel.

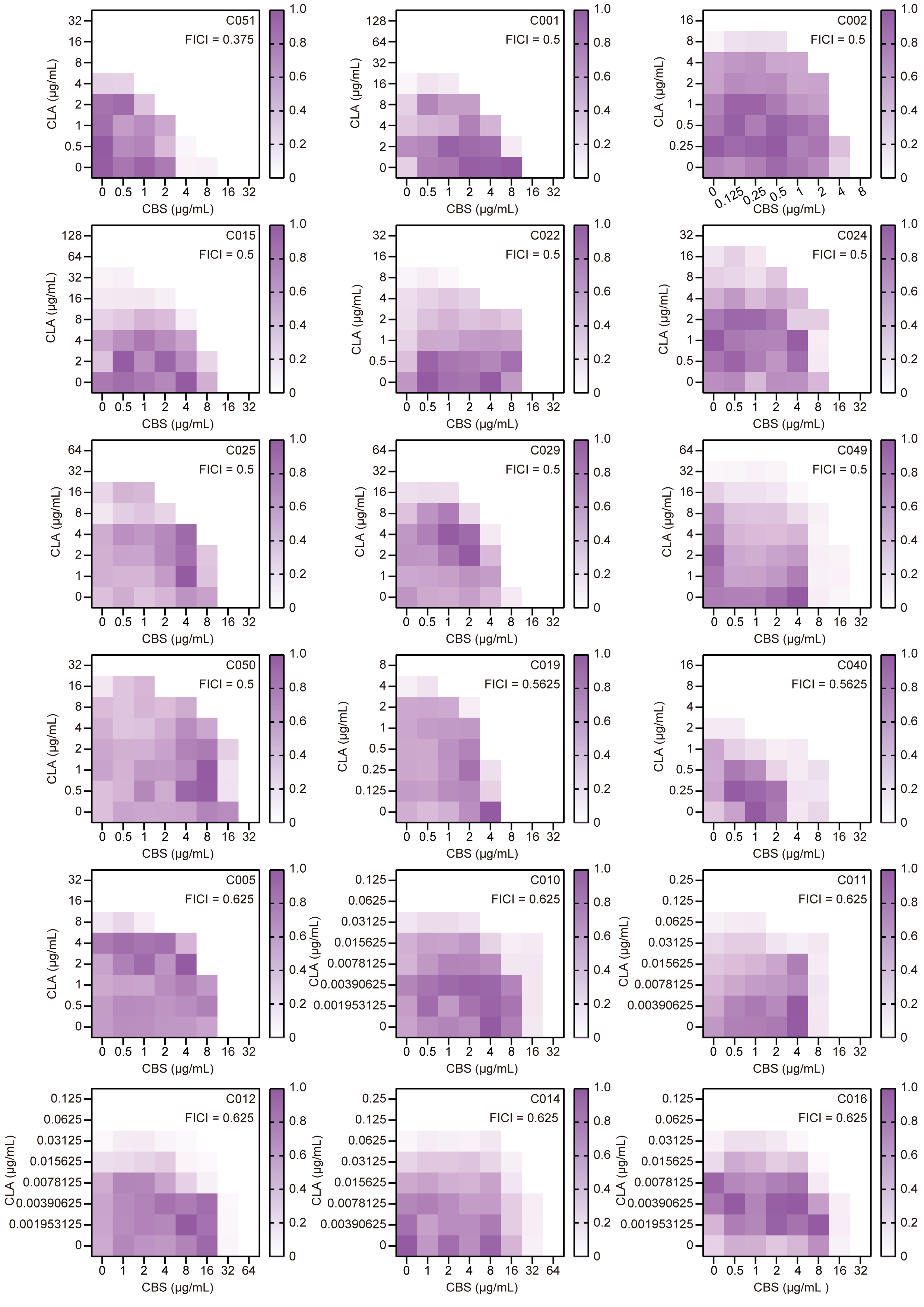

**Figure S3.** Checkerboard assays showing the interaction between CBS and CLA against additional clinical isolates.

Checkerboard microdilution assays were conducted to assess the combined antibacterial activity of CBS and CLA against additional clinical isolates not shown in Figure S2. Drug interactions were interpreted based on the fractional inhibitory concentration index (FICI), with synergy defined as FICI ≤ 0.5 and additive interactions defined as FICI values between 0.5 and 1.0. The isolates presented here indicate predominantly additive or near‑additive interactions between CBS and CLA.

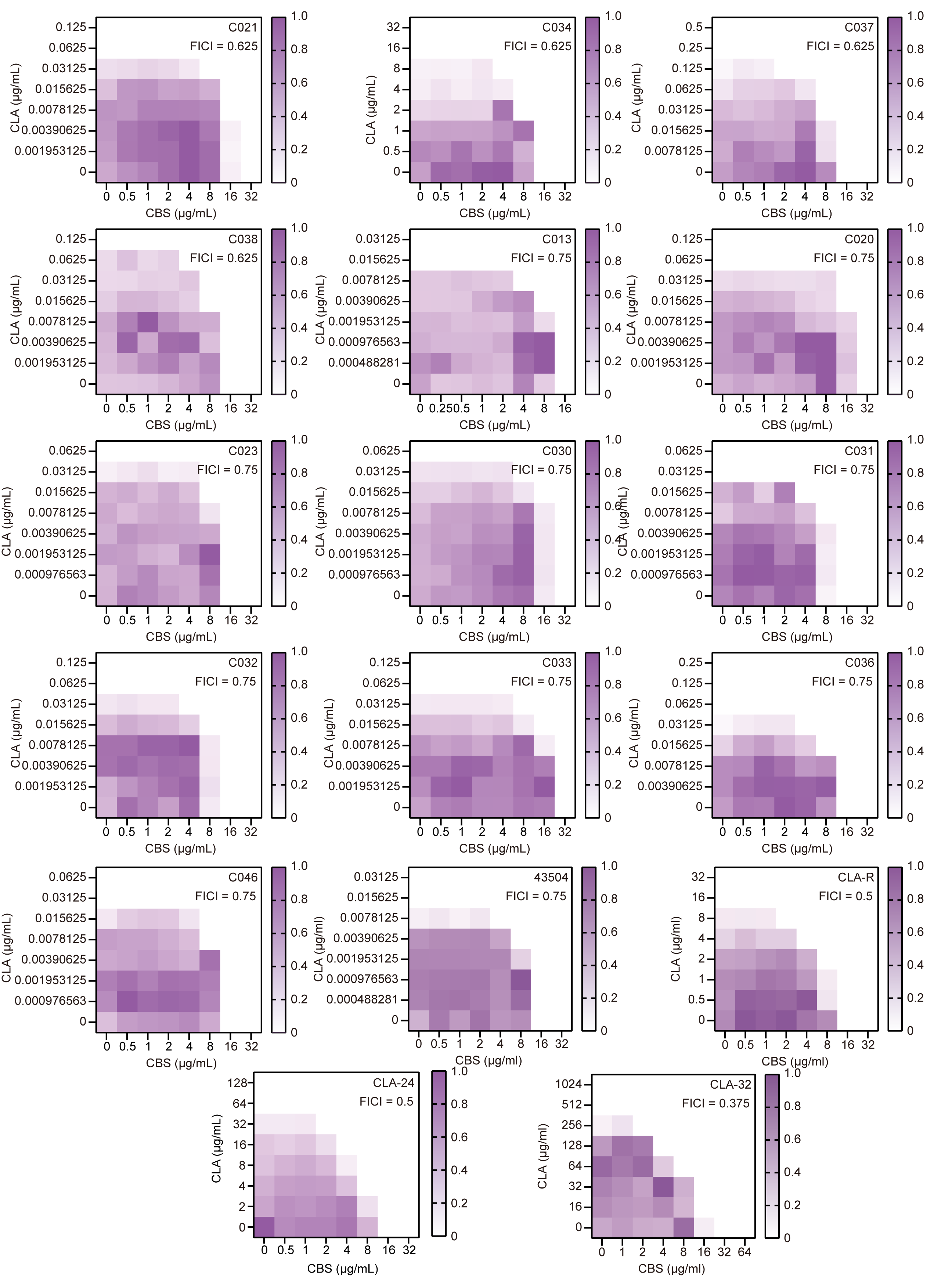

****Figure S4.** Checkerboard assays of CBS and CLA against additional clinical isolates and reference strains.**

**Checkerboard microdilution assays were performed to evaluate the combined antibacterial activity of CBS and CLA against additional clinical isolates and reference strains not shown in Figures S2 and S3. This figure also includes checkerboard assays performed with the reference strain ATCC 43504 and its derivatives exhibiting stepwise increases in CLA resistance following induction at different CLA concentrations. Drug interactions were interpreted using the fractional inhibitory concentration index (FICI), with synergy defined as FICI ≤ 0.5 and additive interactions defined as FICI values between 0.5 and 1.0. The FICI value for each strain is indicated in the upper right corner of each panel. Together with the results shown in Figures S2 and S3, these data indicate that the interaction between CBS and CLA is strain‑dependent, with synergistic or additive effects observed in a subset of strains and predominantly additive or indifferent interactions across a broader panel.**

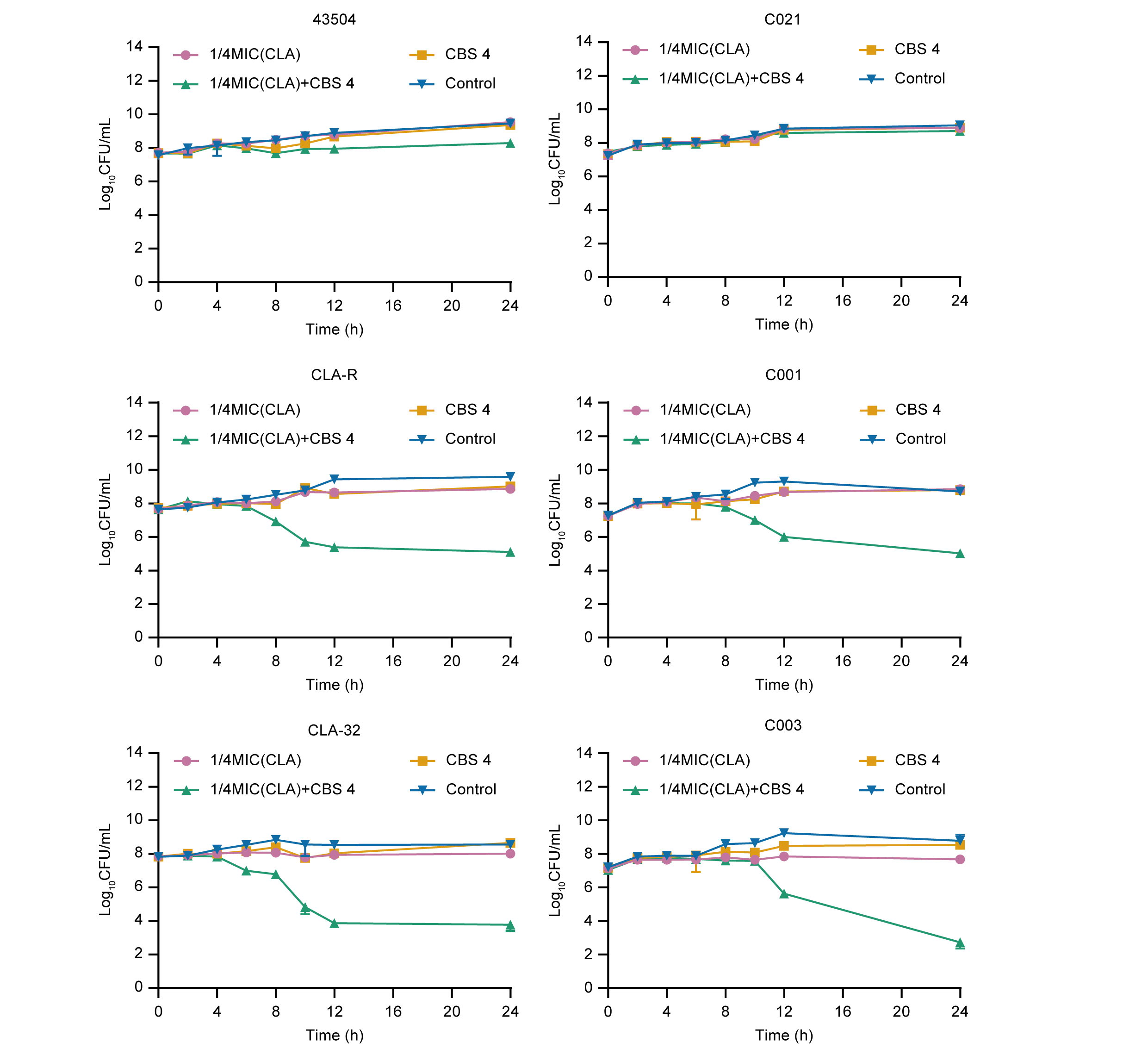

**Figure S5** Time‑kill kinetics of sub‑MIC clarithromycin (CLA) in combination with CBS.

Time‑kill assays were performed in reference strain ATCC 43504, clarithromycin‑resistant derivatives (CLA‑R and CLA‑32), and representative clinical isolates (C001, C003, and C021). Bacterial viability was monitored over 24 h under treatment with CBS alone (4 μg/mL), CLA alone at 1/4 MIC, or the combination of 1/4 MIC of CLA and CBS.

In CLA-susceptible backgrounds (ATCC 43504 and C021), neither single treatments nor the combination caused a significant reduction in CFU counts. In contrast, CLA‑resistant strains exhibited a pronounced time‑dependent decrease in bacterial viability only under combination treatment, while either agent alone remained largely bacteriostatic.

Data are presented as Log_10_CFU/mL (mean ± SD) from at least three independent experiments.

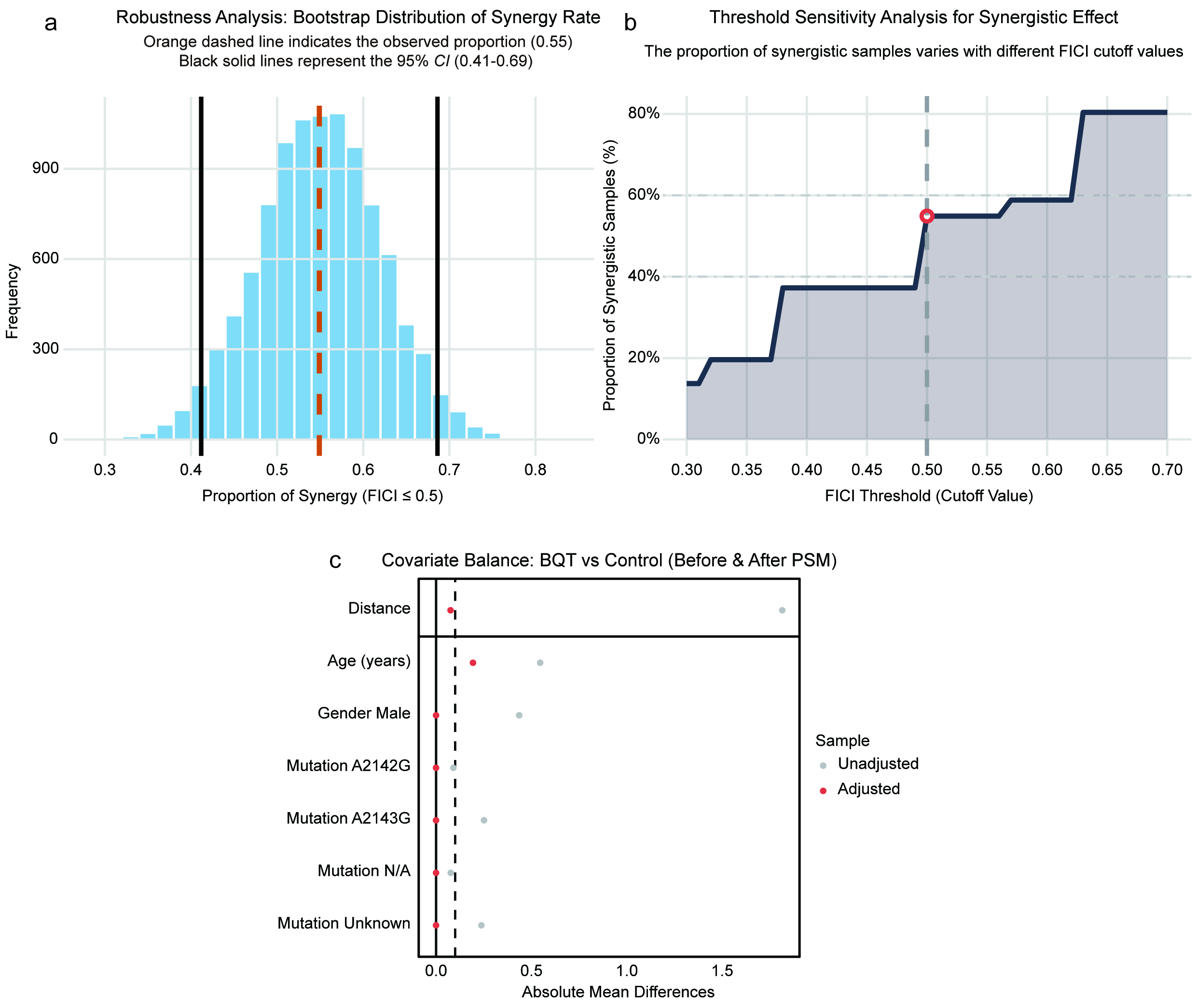

### ****Figure S6.** Covariate balance and robustness analysis of synergistic effects.**

### **(a) Bootstrap robustness analysis. One thousand bootstrap resamples were performed to assess the stability of the observed synergy rate (defined as fractional inhibitory concentration index [FICI] ≤ 0.5). The histogram shows the distribution of synergy proportions across resamples. The orange dashed line represents the observed proportion in the original dataset (0.55), and the black solid lines indicate the 95% bootstrap confidence interval (0.41–0.69). The narrow confidence interval confirms that the association between high-level clarithromycin resistance (MIC ≥ 16 μg/mL) and CBS–CLA synergy is robust and not driven by outlier observations.**

### **(b) Threshold sensitivity analysis across FICI cutoffs. To evaluate whether the observed synergistic effect is dependent on the specific FICI threshold used, the proportion of samples classified as synergistic was calculated across a range of FICI cutoff values (0.30 to 0.70). This analysis demonstrates that the classification remains stable across commonly accepted synergy definitions, supporting the reliability of the identified MIC–synergy relationship.**

### **(c) Covariate balance before and after propensity score matching (PSM). To minimize selection bias in the clinical validation cohort (n = 49), 1:1 nearest-neighbor PSM was performed using caliper = 0.2, with age, sex, and 23S rRNA mutation status as matching covariates. Absolute standardized mean differences (SMDs) for each covariate are shown before (unadjusted, gray) and after (adjusted, red) matching. SMD < 0.1 (vertical dashed line) indicates adequate balance. Following PSM, all covariates achieved balanced distributions between the bismuth-containing quadruple therapy (BQT) and triple therapy (TT) groups, supporting that the observed superiority of BQT in patients with MIC of CLA ≥ 16 μg/mL is not attributable to measurable baseline imbalances.**

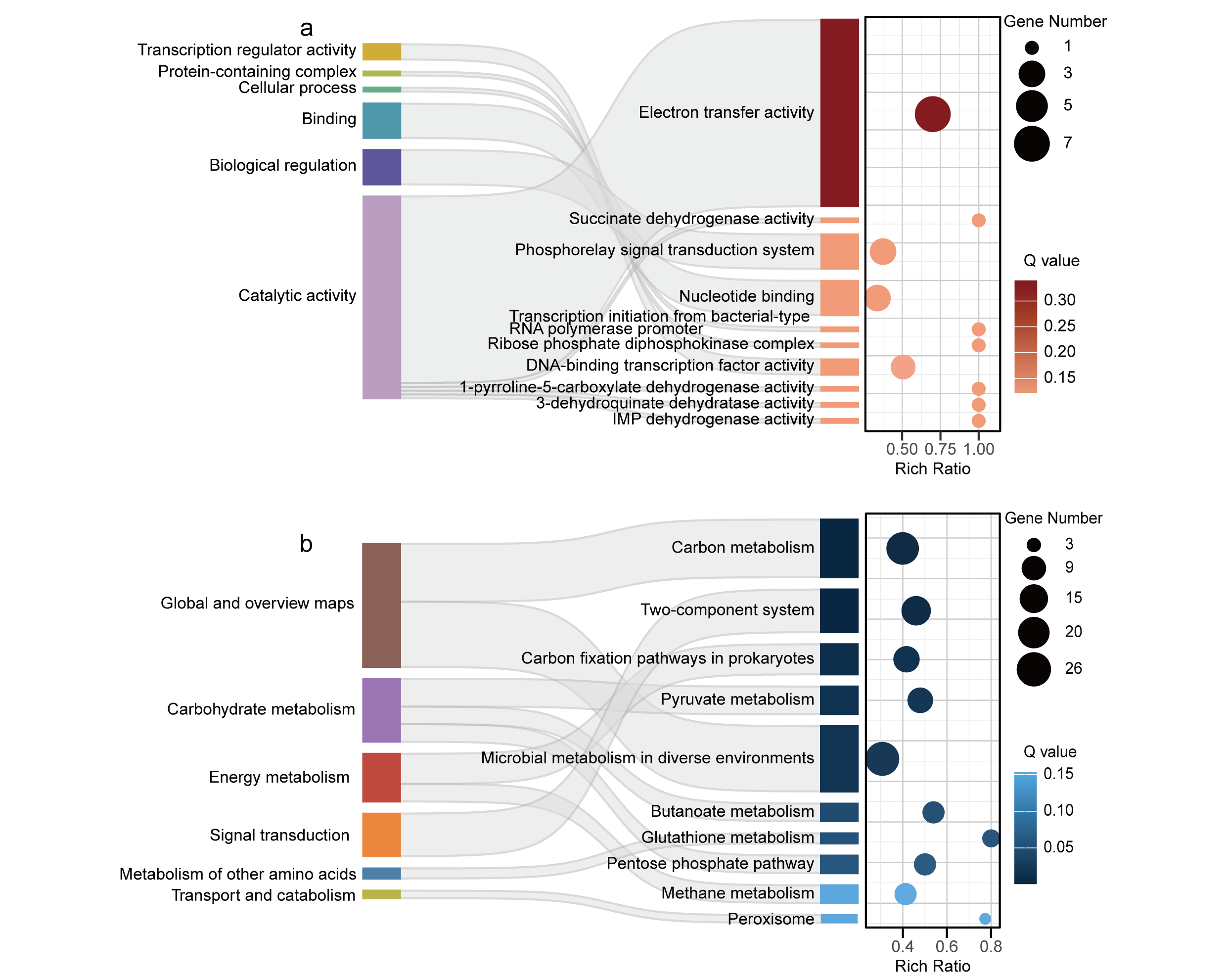

### ****Figure S7.** Functional enrichment analysis of differentially expressed genes upon CBS and CLA treatment**

(a) Gene Ontology (GO) enrichment analysis of differentially expressed genes. Sankey diagram illustrates the relationships between enriched GO categories (left) and specific GO terms (right), including molecular function, biological process, and cellular component. The accompanying bubble plot indicates the richness ratio of each GO term, with bubble size representing the number of associated genes and color denoting the adjusted Q value.

(b) Kyoto Encyclopedia of Genes and Genomes (KEGG) pathway enrichment analysis of differentially expressed genes. Sankey diagram shows the distribution of genes among major functional classes (left) and significantly enriched pathways (right). Bubble plot depicts the enrichment level of each pathway, where bubble size corresponds to gene number and color indicates the Q value.

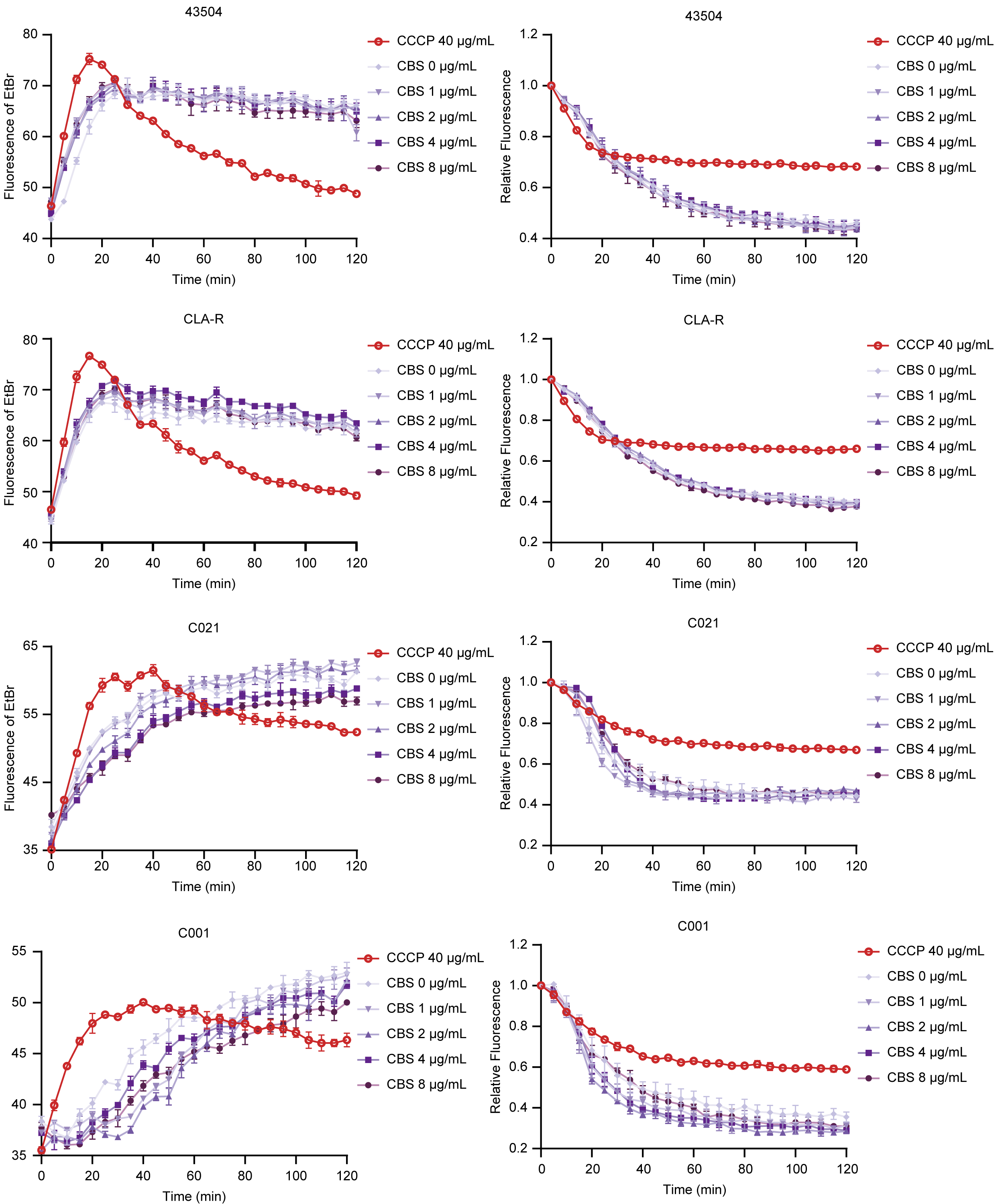

### ****Figure S8.** Evaluation of EtBr uptake and efflux in remaining strains following CBS treatment**

Ethidium bromide (EtBr) uptake (left panels) and efflux (right panels) assays were performed on remaining strains from both the reference strain panel (43504, CLA‑R) and clinical isolate panel (C021, C001).

- ****Uptake assays****: Fluorescence intensity was measured over 120 min to assess EtBr accumulation in the presence of increasing concentrations of CBS (0, 1, 2, 4, 8 μg/mL) or the positive control CCCP (40 μg/mL). CBS treatment did not significantly alter EtBr accumulation compared to untreated controls, indicating no inhibitory effect on EtBr uptake.
- ****Efflux assays****: Relative fluorescence was measured after initial EtBr loading, following the addition of glucose to induce efflux. CCCP treatment partially inhibited efflux, resulting in a sustained higher level of relative fluorescence, while CBS treatment did not suppress efflux activity across all tested concentrations.

These results are consistent with the findings from the main text, confirming that CBS does not modulate EtBr uptake or efflux in these strains, and that CCCP only partially inhibits efflux mechanisms. The strongest synergistic strains and most drug-resistant strains have been presented in the main figures.

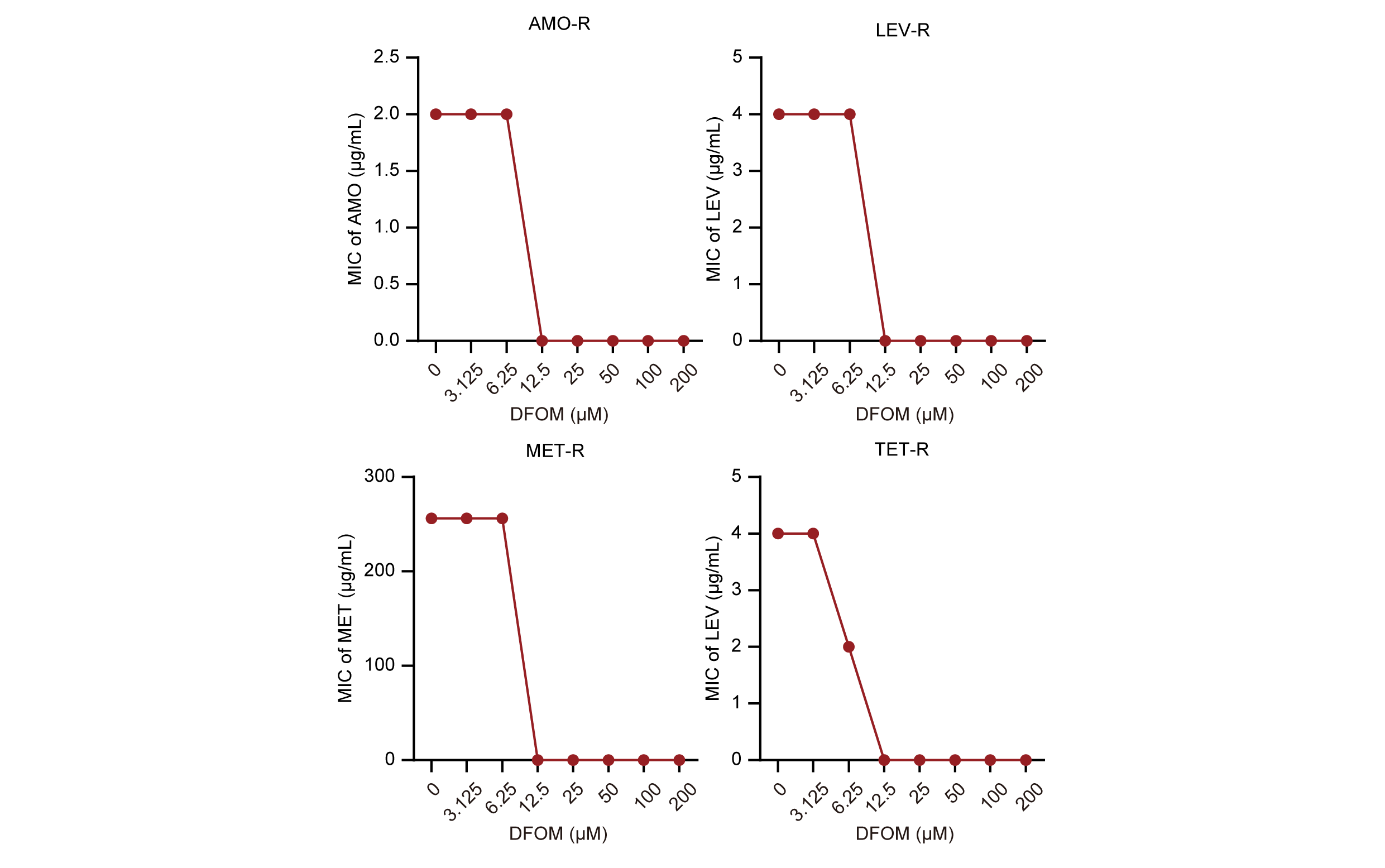

**Figure S9** Iron chelation suppresses antibiotic resistance in resistant backgrounds of ATCC 43504.

Minimum inhibitory concentrations (MICs) of amoxicillin (AMO), levofloxacin (LEV), metronidazole (MET), and tetracycline (TET) were determined for ATCC 43504 and its corresponding resistant derivatives (AMO‑R, LEV‑R, and TET‑R), as well as its intrinsically metronidazole‑resistant background, in the presence of increasing concentrations of the iron chelator deferoxamine mesylate (DFOM). DFOM treatment resulted in a pronounced, dose‑dependent reduction in MICs across all antibiotic backgrounds tested, including intrinsic metronidazole resistance, with near‑complete restoration of susceptibility at higher DFOM concentrations. These results indicate that iron chelation broadly suppresses both intrinsic and acquired antibiotic resistance phenotypes in ATCC 43504.

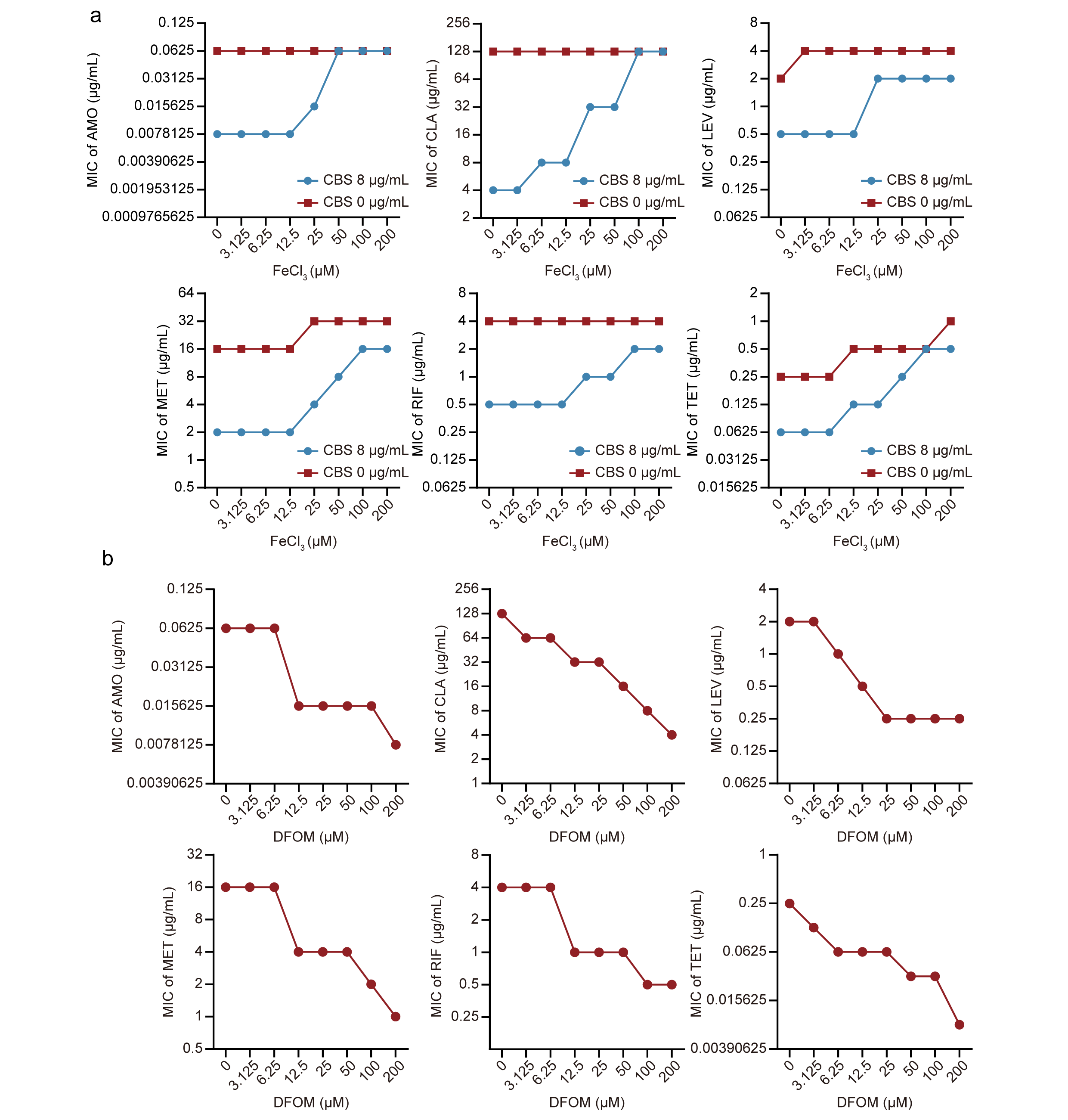

**Figure S9** Iron availability gates the expansion of CBS-mediated antibiotic synergy beyond CLA in a clinical CLA-resistant isolate

The effects of iron supplementation and iron chelation on the synergistic activity of CBS with different antibiotics were evaluated in the clinical CLA‑resistant isolate C003.

**(a)** Minimum inhibitory concentrations (MICs) of amoxicillin (AMO), clarithromycin (CLA), levofloxacin (LEV), metronidazole (MET), rifampicin (RIF), and tetracycline (TET) were determined in the presence or absence of CBS (8 μg/mL) under increasing concentrations of ferric chloride (FeCl₃). Iron supplementation progressively antagonized the CBS-mediated reduction in MICs, indicating that excess iron compromises the synergistic effects of CBS with multiple antibiotics.

**(b)** MICs of the same antibiotics were measured in the presence of increasing concentrations of deferoxamine mesylate (DFOM). Iron chelation resulted in a dose-dependent decrease in MICs across all tested antibiotics, consistent with enhanced antibiotic susceptibility under iron-limited conditions.

Together, these results demonstrate that CBS exhibits synergistic activity with multiple antibiotics in the CLA‑resistant clinical isolate C003, and that this synergy is negatively regulated by iron availability.
