## Supplementary material for "High-level clarithromycin resistance: a metabolic vulnerability exploited by bismuth in *Helicobacter pylori*": Table S1-S6

**Supplementary Tables**

**Table S1. Baseline characteristics of the study population**

| **Baseline** | **Follow-up**  **(n=1844)**  **(95%CI)** | **Lost-follow-up (n=2766)**  **(95%CI)** | **χ^2^** | ***p*-Value** |
| --- | --- | --- | --- | --- |
| **Sex** |  |  |  |  |
| Male | 57.7% (1064/1844)  (55.4-60.0%) | 56.1% (1552/2766)  (54.2-58.0%) | 1.14 | 0.286 |
| Female | 42.3% (780/1844)  (40.0-44.6%) | 43.9% (1214/2766)  (42.0-45.8%) |  |  |
| **Age (years)** | 9.3±3.2 | 9.9±3.2 |  | 0.060 |
| **Endoscopic performance** |  |  |  |  |
| Non-ulcerative erosion lesion | 80.5% (1484/1844)  (78.6-82.3%) | 84.2% (2329/2766)  (82.8-85.5%) | 11.38 | 0.001** |
| Ulcerative erosion lesion | 19.5% (360/1844)  (17.7-21.4%) | 15.8% (437/2766)  (14.5-17.2%) |  |  |
| **Culture positive rate** | 29.2% (313/1073)  (26.5-32.0%) | 34.4% (430/1251)  (31.7-37.1%) | 7.19 | 0.007** |
| **Antibiotic resistance** | n=176  (176/313, 56.2%)  (50.5-61.8%) | n=196  (196/430, 45.6%)  (40.8-50.4%) | 8.22 | 0.004** |
| AMO | 5.1% (9/176)  (2.4-9.5%) | 3.6% (7/196)  (1.4-7.2%) | 0.54 | 0.464 |
| CLA | 39.2% (69/176)  (31.9-46.8%) | 43.9% (86/196)  (36.8-51.1%) | 2.12 | 0.146 |
| LEV | 1.1% (2/176)  (0.1-4.0%) | 1.0% (2/196)  (0.1-3.6%) | 0.01 | 1.000 |
| MET | 88.1% (155/176)  (82.3-92.5%) | 89.8% (176/196)  (84.7-93.7%) | 0.28 | 0.595 |
| **Bismuth-containing quadruple therapy** | n=995  (995/1844, 54.0% )  (51.7-56.3%) | n=1699  (1699/2766, 61.4%)  (59.6-63.2%) | 25.39 | < 0.0001**** |
| CBS+AMO+MET+PPI | 55.9% (556/995)  (52.7-59.0%) | 60.6% (1030/1699)  (58.3-63.0%) | 5.26 | 0.022 |
| CBS+CLA+AMO+PPI | 14.5% (144/995)  (12.3-16.8%) | 15.8% (268/1699)  (14.1-17.6%) | 0.82 | 0.365 |
| CBS+CLA+MET+PPI | 29.6% (295/995)  (26.8-32.6%) | 23.6% (401/1699)  (21.6-25.7%) | 11.97 | 0.001** |
| **Concomitant therapy：**  AMO+MET+CLA+PPI | 2.4% (44/1844)  (1.7-3.2%) | 2.4% (65/2766)  (1.8-3.0%) | 0.02 | 0.888 |
| **Triple therapy** | n=805  (805/1844, 43.6%)  (41.4-46.0%) | n=1002  (1002/2766, 36.2%)  (34.4-38.0%) | 25.62 | < 0.0001**** |
| AMO+MET+PPI | 7.5% (60/805)  (5.7-9.5%) | 6.5% (65/1002)  (5.0-8.2%) | 0.65 | 0.421 |
| AMO+CLA+PPI | 76.0% (612/805)  (72.9-78.9%) | 77.8% (780/1002)  (75.1-80.4%) | 0.84 | 0.361 |
| CLA+MET+PPI | 16.5% (133/805)  (14.0-19.3%) | 15.7% (157/10 02)  (13.5-18.1%) | 0.24 | 0.623 |

Categorical variables are presented as counts and percentages, with 95% confidence intervals (CI). Continuous variables are presented as mean ± standard deviation (SD). Comparisons between groups were performed using the *χ*^2^ test or Fisher’s exact test, as appropriate.

Antibiotic resistance was defined according to EUCAST breakpoints (version 13.0).

** *p* < 0.01; **** *p* < 0.001.

**Table S2. Comparison of eradication efficacy among bismuth-containing quadruple therapy, concomitant therapy, and triple therapy**

| **Therapy** | **Eradication rate (95%CI)** | ***χ*^2^** | ***p*-Value** |
| --- | --- | --- | --- |
| **Bismuth-containing quadruple therapy** | 87.1% (867/995) (84.9-89.2%) | 58.0^(1)^ | <0.0001****^(1)^ |
| CBS+AMO+MET+PPI | 91.9% (511/556) (89.3-94.0%) |  | < 0.0001**** |
| CBS+CLA+AMO+PPI | 85.4% (123/144) (78.6-90.7%) |  |  |
| CBS+CLA+MET+PPI | 81.8% (233/285) (76.8-86.1%) |  |  |
| **Concomitant therapy**  AMO+MET+CLA+PPI | 81.8% (36/44) (67.3-91.8%) |  |  |
| **Triple therapy** | 72.9% (587/805) (69.7-76.0%) |  |  |
| AMO+MET+PPI | 73.3% (44/60) (60.3-83.9%) | 4.58 | 0.101 |
| AMO+CLA+PPI | 74.5% (456/612) (70.9-77.9%) |  |  |
| CLA+MET+PPI | 65.4% (87/133) (56.7-73.4%) |  |  |

Categorical variables are presented as counts and percentages, along with their 95% CI. *****p* < 0.0001.

**Table S3. Demographic and clinical characteristics of *H. pylori* isolates stratified by antibiotic resistance profiles**

| **Therapy** | **Eradication Rate (95%CI)** | ***χ*^2^** | ***p*-Value** |
| --- | --- | --- | --- |
| **AMO resistance** | 44.4% (4/9) (13.7-78.8%) | 1.40 |  |
| **Bismuth-containing quadruple therapy** | 100.0% (1/1) (2.5-100%) |  |  |
| CBS+CLA+MET+PPI | 100.0% (1/1) (2.5-100%) |  |  |
| **Triple therapy** | 37.5% (3/8) (8.5-75.5%) |  |  |
| AMO+CLA+PPI | 28.6% (2/7) (3.7-71.0%) |  |  |
| CLA+MET+PPI | 100.0% (1/1) (2.5-100%) |  |  |
| **LEV resistance** | 100.0% (2/2) (15.8-100%) |  |  |
| **Bismuth-containing quadruple therapy** | 100.0% (1/1) (2.5-100%) |  |  |
| CBS+CLA+MET+PPI | 100.0% (1/1) (2.5-100%) |  |  |
| **Triple therapy** | 100.0% (1/1) (2.5-100%) |  |  |
| AMO+CLA+PPI | 100.0% (1/1) (2.5-100%) |  |  |
| **CLA resistance** | 73.9% (51/69) (62.2%–83.4%) |  |  |
| **BQT** | 93.1% (27/29) (77.2-99.2%) | 0.57 | 1.000 |
| CBS+AMO+MET+PPI | 92.9% (13/14) (66.1-99.8%) |  |  |
| CBS+CLA+AMO+PPI | 100.0% (4/4) (39.8-100%) |  |  |
| CBS+CLA+MET+PPI | 90.9% (10/11) (58.7-99.8%) |  |  |
| **CT**  AMO+MET+CLA+PPI | 100.0% (2/2) (15.8-100%) |  |  |
| **TT** | 57.9% (22/38) (41.3%–73.2%) | 10.41^#^ | 0.001**^#^ |
| AMO+MET+PPI | 100.0% (2/2) (15.8–100.0%%) |  |  |
| AMO+CLA+PPI | 67.7% (21/31) (48.6–83.3%) |  |  |
| CLA+MET+PPI | 60.0% (3/5) (14.6–94.7%) |  |  |
| **MET resistance** | 77.4% (120/155) (70.0-83.7%) |  |  |
| **Bismuth-containing quadruple therapy** | 80.0% (56/70) (68.7-88.6%) | 2.317 | 0.247 |
| CBS+AMO+MET+PPI | 88.6% (31/35) (73.3-96.8%) |  |  |
| CBS+CLA+AMO+PPI | 87.5% (7/8) (47.3-99.7%) |  |  |
| CBS+CLA+MET+PPI | 66.7% (18/27) (46.0-83.5%) |  |  |
| **Concomitant therapy**  AMO+MET+CLA+PPI | 50.0% (2/2) (6.8-93.2%) |  |  |
| **Triple therapy** | 73.8% (59/80) (62.7-83.0%) | 0.81^#^ | 0.367^#^ |
| AMO+MET+PPI | 80.0% (4/5) (28.4-99.5%) |  |  |
| AMO+CLA+PPI | 72.5% (50/69) (60.4-82.5%) |  |  |
| CLA+MET+PPI | 83.3% (5/6) (35.9-99.6%) |  |  |

Categorical variables are presented as counts and percentages, along with their 95% CI. Fisher’s exact test was used when expected cell counts were <5. Given the limited number of patients receiving concomitant therapy in resistance-stratified analyses, pairwise comparisons were performed exploratorily to assess differential treatment effects.

\# Exploratory pairwise comparison between bismuth-containing quadruple therapy and standard triple therapy. * *p* < 0.05.

**Table S4. Stratification of clinical *Helicobacter pylori* isolates by MIC of CLA reveals a tripartite response to CBS–CLA combination: susceptibility (≤ 0.25 µg/mL), intermediate resistance (0.5 – 8 µg/mL), and high-level resistance (≥ 16 µg/mL) associated with synergy (FICI ≤ 0.5) and successful eradication**

| **Sample** | **MIC of CBS** | **MIC of CLA** | **MIC of CBS/CLA** | **MIC of CLA/CBS** | **FICI** | **Effect** | **Point mutation** | **Therapy** | **Eradic**  **ation** |
| --- | --- | --- | --- | --- | --- | --- | --- | --- | --- |
| C001 | 16 | 32 | 4 | 8 | 0.5 | Synergy | A2143G | BQT | Yes |
| C002 | 8 | 16 | 2 | 4 | 0.5 | Synergy | A2143G | BQT | Yes |
| C003 | 32 | 128 | 2 | 16 | 0.1875 | Synergy | A2143G | BQT | Yes |
| C004 | 32 | 64 | 4 | 16 | 0.375 | Synergy | A2143G | N/A | None |
| C005 | 16 | 16 | 8 | 2 | 0.625 | Additive | A2143G | BQT | Yes |
| C006 | 32 | 64 | 4 | 8 | 0.25 | Synergy | A2143G | TT | No |
| C007 | 32 | 128 | 4 | 16 | 0.25 | Synergy | A2143G | BQT | Yes |
| C008 | 64 | 64 | 16 | 8 | 0.375 | Synergy | A2143G | BQT | Yes |
| C009 | 32 | 0.125 | 16 | 0.03125 | 0.75 | Additive |  | BQT | Yes |
| C010 | 32 | 0.0625 | 4 | 0.03125 | 0.625 | Additive |  | TT | Yes |
| C011 | 16 | 0.125 | 2 | 0.0625 | 0.625 | Additive |  | TT | Yes |
| C012 | 64 | 0.0625 | 32 | 0.00781 | 0.625 | Additive |  | TT | Yes |
| C013 | 16 | 0.015625 | 8 | 0.00391 | 0.75 | Additive |  | N/A | None |
| C014 | 64 | 0.125 | 32 | 0.015625 | 0.625 | Additive |  | TT | Yes |
| C015 | 16 | 64 | 4 | 16 | 0.5 | Synergy | A2143G | BQT | Yes |
| C016 | 32 | 0.0625 | 16 | 0.00781 | 0.625 | Additive |  | BQT | Yes |
| C017 | 32 | 256 | 8 | 32 | 0.375 | Synergy | A2143G | TT | No |
| C018 | 32 | 256 | 4 | 32 | 0.25 | Synergy | A2143G | BQT | Yes |
| C019 | 8 | 8 | 4 | 0.5 | 0.5625 | Additive | N/A | BQT | Yes |
| C020 | 32 | 0.0625 | 16 | 0.015625 | 0.75 | Additive |  | BQT | Yes |
| C021 | 32 | 0.0625 | 16 | 0.00781 | 0.625 | Additive |  | BQT | Yes |
| C022 | 16 | 16 | 4 | 4 | 0.5 | Synergy | A2143G | BQT | Yes |
| C023 | 16 | 0.0625 | 8 | 0.015625 | 0.75 | Additive |  | BQT | Yes |
| C024 | 16 | 32 | 4 | 8 | 0.5 | Synergy | A2143G | BQT | Yes |
| C025 | 16 | 32 | 4 | 8 | 0.5 | Synergy | A2143G | BQT | Yes |
| C026 | 32 | 32 | 8 | 4 | 0.375 | Synergy | A2143G | BQT | Yes |
| C027 | 32 | 512 | 4 | 128 | 0.375 | Synergy | A2142G | TT | No |
| C028 | 32 | 256 | 4 | 16 | 0.1875 | Synergy | A2143G | BQT | Yes |
| C029 | 16 | 32 | 4 | 8 | 0.5 | Synergy | A2143G | BQT | Yes |
| C030 | 32 | 0.0625 | 16 | 0.015625 | 0.75 | Additive |  | BQT | Yes |
| C031 | 16 | 0.03125 | 8 | 0.015625 | 0.75 | Additive |  | BQT | Yes |
| C032 | 16 | 0.0625 | 8 | 0.015625 | 0.75 | Additive |  | TT | No |
| C033 | 32 | 0.0625 | 16 | 0.015625 | 0.75 | Additive |  | BQT | Yes |
| C034 | 16 | 16 | 8 | 2 | 0.625 | Additive | A2143G | BQT | No |
| C035 | 64 | 128 | 4 | 32 | 0.3125 | Synergy | A2143G | BQT | Yes |
| C036 | 16 | 0.0625 | 8 | 0.015625 | 0.75 | Additive |  | BQT | Yes |
| C037 | 16 | 0.25 | 8 | 0.03125 | 0.625 | Additive |  | BQT | Yes |
| C038 | 16 | 0.125 | 8 | 0.015625 | 0.625 | Additive |  | BQT | Yes |
| C039 | 32 | 128 | 4 | 32 | 0.375 | Synergy | A2143G | BQT | Yes |
| C040 | 16 | 4 | 1 | 2 | 0.5625 | Additive | A2143G | BQT | Yes |
| C041 | 64 | 64 | 4 | 16 | 0.3125 | Synergy | A2143G | BQT | Yes |
| C042 | 16 | 512 | 2 | 16 | 0.15625 | Synergy | N/A | BQT | Yes |
| C043 | 16 | 0.25 | 2 | 0.125 | 0.625 | Additive |  | BQT | Yes |
| C044 | 16 | 32 | 4 | 4 | 0.375 | Synergy | A2143G | BQT | Yes |
| C045 | 16 | 16 | 4 | 2 | 0.375 | Synergy | A2143G | BQT | Yes |
| C046 | 16 | 0.03125 | 8 | 0.00781 | 0.75 | Additive |  | BQT | Yes |
| C047 | 64 | 256 | 16 | 16 | 0.3125 | Synergy | A2143G | BQT | Yes |
| C048 | 16 | 64 | 2 | 8 | 0.25 | Synergy | A2143G | BQT | Yes |
| C049 | 32 | 64 | 8 | 16 | 0.5 | Synergy | A2143G | BQT | Yes |
| C050 | 32 | 32 | 8 | 8 | 0.5 | Synergy | N/A | BQT | Yes |
| C051 | 16 | 8 | 2 | 2 | 0.375 | Synergy | A2143G | TT | Yes |
| 43504 | 16 | 0.015625 | 8 | 0.00391 | 0.75 | Additive |  | N/A | N/A |
| CLA-R | 16 | 16 | 4 | 4 | 0.5 | Synergy | A2143G | N/A | N/A |
| CLA-24 | 16 | 64 | 4 | 16 | 0.5 | Synergy | A2143G | N/A | N/A |
| CLA-32 | 32 | 512 | 4 | 128 | 0.375 | Synergy | A2143G | N/A | N/A |

Abbreviations: CBS, colloidal bismuth subcitrate; CLA, clarithromycin; MIC, minimum inhibitory concentration; FICI, fractional inhibitory concentration index; BQT, bismuth-containing quadruple therapy; TT, Triple therapy; N/A, Not Applicable.

FICI was calculated as (MIC of CBS in combination / MIC of CBS alone) + (MIC of CLA in combination / MIC of CLA alone). MICs are expressed in μg/mL.

Point mutations refer to substitutions in the 23S rRNA gene associated with CLA resistance.

Interactions were classified as synergistic (FICI ≤ 0.5) or additive (0.5 < FICI ≤ 1.0).

**Table S5. Diagnostic performance of MIC thresholds for predicting synergy (FICI ≤0.5) with clarithromycin (CLA).**

| **MIC of CLA Cut-off (µg/mL)** | **Sensitivity, %** | **Specificity, %** | **Youden’s Index** |
| --- | --- | --- | --- |
| >512 | 0.0 | 100.0 | 0.000 |
| 512 | 7.1 | 100.0 | 0.071 |
| 256 | 21.4 | 100.0 | 0.214 |
| 128 | 35.7 | 100.0 | 0.357 |
| 64 | 60.7 | 100.0 | 0.607 |
| 32 | 85.7 | 100.0 | 0.857 |
| **16** | **96.4** | **91.3** | **0.877** |
| 8 | 100.0 | 87.0 | 0.870 |
| 4 | 100.0 | 82.6 | 0.826 |
| 0.25 | 100.0 | 73.9 | 0.739 |
| 0.125 | 100.0 | 56.5 | 0.565 |
| 0.0625 | 100.0 | 13.0 | 0.130 |
| 0.03125 | 100.0 | 4.3 | 0.043 |
| 0.015625 | 100.0 | 0.0 | 0.000 |
| <0.015625 | 100.0 | 0.0 | 0.000 |

FICI, fractional inhibitory concentration index. The gold standard for synergistic interaction was defined as FICI ≤ 0.5. The area under the receiver operating characteristic curve (AUC) was 0.992. The optimal cut-off (MIC ≥ 16 mg/L) is highlighted in bold, based on the maximum Youden’s Index.

**Table S6. MICs of CLA and FICI values of CBS combinations in reference strain and clinical isolate panels**

| **MIC**  **(FICI)** | **Reference strain panel** | | | **Clinical isolate panel** | | |
| --- | --- | --- | --- | --- | --- | --- |
|  | **43504** | **CLA-R** | **CLA-32** | **C021** | **C001** | **C003** |
| CLA | 0.015625  (0.75) | 16  (0.5) | 512  (0.375) | 0.0625  (0.625) | 32  (0.5) | 128  (0.1875) |
| AMO | 0.0625  (0.75) | 0.0625  (0.625) | 0.25  (0.5) | 0.125  (1) | 0.5  (0.75) | 0.0625  (0.375) |
| LEV | 0.25  (2) | 1  (0.625) | 1  (0.515625) | 8  (2) | 0.25  (1) | 2  (0.5) |
| MET | 256  (0.75) | 256  (0.625) | 512  (0.5) | 256  (0.5) | 32  (0.53125) | 16  (0.375) |
| RIF | 0.25  (0.75) | 0.5  (0.625) | 2  (0.25) | 8  (0.5625) | 8  (0.375) | 4  (0.375) |
| TET | 0.125  (2) | 2  (1) | 2  (0.75) | 2  (0.75) | 0.5  (0.75) | 0.25  (0.5) |

FICI ≤ 0.5, synergy; 0.5 < FICI ≤ 1, additive; 1 < FICI ≤ 2, indifferent; FICI > 2, antagonism.

MICs are expressed in μg/mL.
